# Utilizing multimodal AI to improve genetic analyses of cardiovascular traits

**DOI:** 10.1101/2024.03.19.24304547

**Authors:** Yuchen Zhou, Justin Cosentino, Taedong Yun, Mahantesh I. Biradar, Jacqueline Shreibati, Dongbing Lai, Tae-Hwi Schwantes-An, Robert Luben, Zachary McCaw, Jorgen Engmann, Rui Providencia, Amand Floriaan Schmidt, Patricia Munroe, Howard Yang, Andrew Carroll, Anthony P. Khawaja, Cory Y. McLean, Babak Behsaz, Farhad Hormozdiari

## Abstract

Electronic health records, biobanks, and wearable biosensors contain multiple high-dimensional clinical data (HDCD) modalities (e.g., ECG, Photoplethysmography (PPG), and MRI) for each individual. Access to multimodal HDCD provides a unique opportunity for genetic studies of complex traits because different modalities relevant to a single physiological system (e.g., circulatory system) encode complementary and overlapping information. We propose a novel multimodal deep learning method, M-REGLE, for discovering genetic associations from a joint representation of multiple complementary HDCD modalities. We showcase the effectiveness of this model by applying it to several cardiovascular modalities. M-REGLE jointly learns a lower representation (i.e., latent factors) of multimodal HDCD using a convolutional variational autoencoder, performs genome wide association studies (GWAS) on each latent factor, then combines the results to study the genetics of the underlying system. To validate the advantages of M-REGLE and multimodal learning, we apply it to common cardiovascular modalities (PPG and ECG), and compare its results to unimodal learning methods in which representations are learned from each data modality separately, but the downstream genetic analyses are performed on the combined unimodal representations. M-REGLE identifies 19.3% more loci on the 12-lead ECG dataset, 13.0% more loci on the ECG lead I + PPG dataset, and its genetic risk score significantly outperforms the unimodal risk score at predicting cardiac phenotypes, such as atrial fibrillation (Afib), in multiple biobanks.

## 1 Introduction

High-dimensional clinical data (HDCD; e.g., ECG, PPG, MRI) are valuable assets for clinical diagnosis, treatment, and prognosis, and provide a unique opportunity for studying the genetic basis of complex traits [1–9]. Recent technological progress in electronic health record (EHR) systems enables access to multiple HDCD modalities per individual [10–12]. When multiple clinical modalities pertain to a single organ system (e.g., circulatory system) or disease process (e.g., cardiovascular diseases), these modalities may encode complementary information. To maximize information for genetic association analyses, such as genome-wide association studies (GWAS), we propose to extract information from these modalities via a joint representation model.

In the past few years, a large body of work has utilized multimodal learning in medicine and biology. However, to the best of our knowledge, these have not been applied to genetic discovery and risk prediction. The existing efforts in medicine range from disease progression prediction [13], disease subtyping [14, 15], healthcare applications [12], disease diagnoses [16], and recently, general medical capabilities utilizing foundation models [17–20]. Furthermore, many biological datasets have utilized multimodal data such as chromatin accessibility, gene expression, protein, and others to improve the biological understanding of cell structure, subpopulations, and physiology [21–25]. In this work, we expand these recent efforts into the field of genomics. Recently, a new method has been developed to perform GWAS on HDCD, called REpresentation learning for Genetic Discovery on Low-dimensional Embeddings (REGLE) [8]. REGLE uses convolutional variational autoencoders (VAEs) to compute a *non-linear, low-dimensional, disentangled embedding* of the data without clinical labels. Although REGLE’s performance in genetic analysis and down-stream polygenic risk scoring (PRS) surpasses prior methods [8], the utility of REGLE is limited to one data modality.

Different clinical modalities can include both complementary and overlapping information. When we model each modality separately using unimodal representation learning, as in the case of REGLE, we cannot leverage these properties to the fullest. We hypothesized that jointly modeling multiple complementary modalities would aid in learning more informative representations, boost the biological signal, and mitigate noise, thus improving genetic discovery and polygenic risk prediction. Furthermore, the shared information among related modalities is captured more efficiently in the joint embedding space, as opposed to having duplicated information in separate representations for each modality. This reserves more modeling capacity for identifying independent biological signals that are unique to each modality. Incorporating such signals can lead to new genetic discoveries.

In this work, we proposed a new method, called M-REGLE (Multimodal REGLE), where we extended REGLE to incorporate multimodal data. In M-REGLE, we first conducted a test to check if two modalities contain complementary information. If they do, we jointly learned a lower-dimensional representation (i.e., latent factors) of the multimodal HDCD using convolutional VAEs [26] (i.e., use early fusion), orthogonalized the latent space, performed GWAS on each orthogonalized latent factor, and finally combined them to study the genetics of the underlying system. We validated the advantages of multimodal learning via M-REGLE by comparing it to unimodal learning in which representations were learned on each data modality separately, and downstream analyses were performed on the concatenation of all representations (i.e., late fusion). We call this unimodal REGLE (U-REGLE). Two sets of cardiovascular modalities in the UK Biobank dataset were used in the experiments: 12 leads of ECG as separate modalities, and the lead I of ECG and PPG (the modalities prevalent in modern smart watches). In both sets of multimodal data, M-REGLE not only detected more novel loci, but also significantly improved genetic risk scoring for cardiac phenotypes, such as atrial fibrillation (Afib).

Our main contributions are as follows: 1) We utilized 12-lead ECG and PPG data in UK Biobank to demonstrate the utility of multimodal modeling for genetic analysis and PRS. Most of the popular smart wearable devices contain sensors which allow users to record their ECG (equivalent to the lead I ECG) and PPG. Our experiments demonstrated that M-REGLE will enable researchers and users to optimally leverage this newly surging data for genetic discovery. 2) We observed that M-REGLE improved genomics discovery over existing methods for 12-lead ECG dataset and ECG lead I + PPG dataset for three metrics: Number of loci (19.3% more loci for 12-lead ECG and 13.0% more loci on the ECG lead I + PPG), number of hits (31.0% more loci for 12-lead ECG and 15.7% more loci on the ECG lead I + PPG), and expected chi-square statistics as measure of power (22.0% higher *E*[*χ*^2^] for 12-lead ECG and 16.4% higher *E*[*χ*^2^] for ECG lead I + PPG). 3) We observed that PRS obtained from M-REGLE for 12-lead ECG significantly outperformed the state of the art method on cardiovascular traits and in particular atrial fibrillation (Afib) in UK Biobank. These results were independently validated in Indiana Biobank, EPIC-Norfolk datasets, and British Women’s Heart and Health Study.

## 2 Results

### 2.1 Overview of M-REGLE

M-REGLE (Multimodal REpresentation learning for Genetic discovery on Low-dimensional Embeddings) learns the low-dimensional embeddings of multiple modalities in a joint model to improve genetic analyses compared to learning embeddings from each modality individually then combining them for the genetic analyses. It is more effective in the presence of complementary and overlapping information across modalities. There are three steps in M-REGLE: 1) jointly learning a *general* non-linear, low dimensional, disentangled representation from multimodal HDCD, 2) obtaining completely uncorrelated embeddings by computing principal components (PCs) of the embeddings, and 3) performing GWAS on each PC, combining them, then performing downstream analyses on combined summary statistics (Figure 1a).

**Figure 1:**
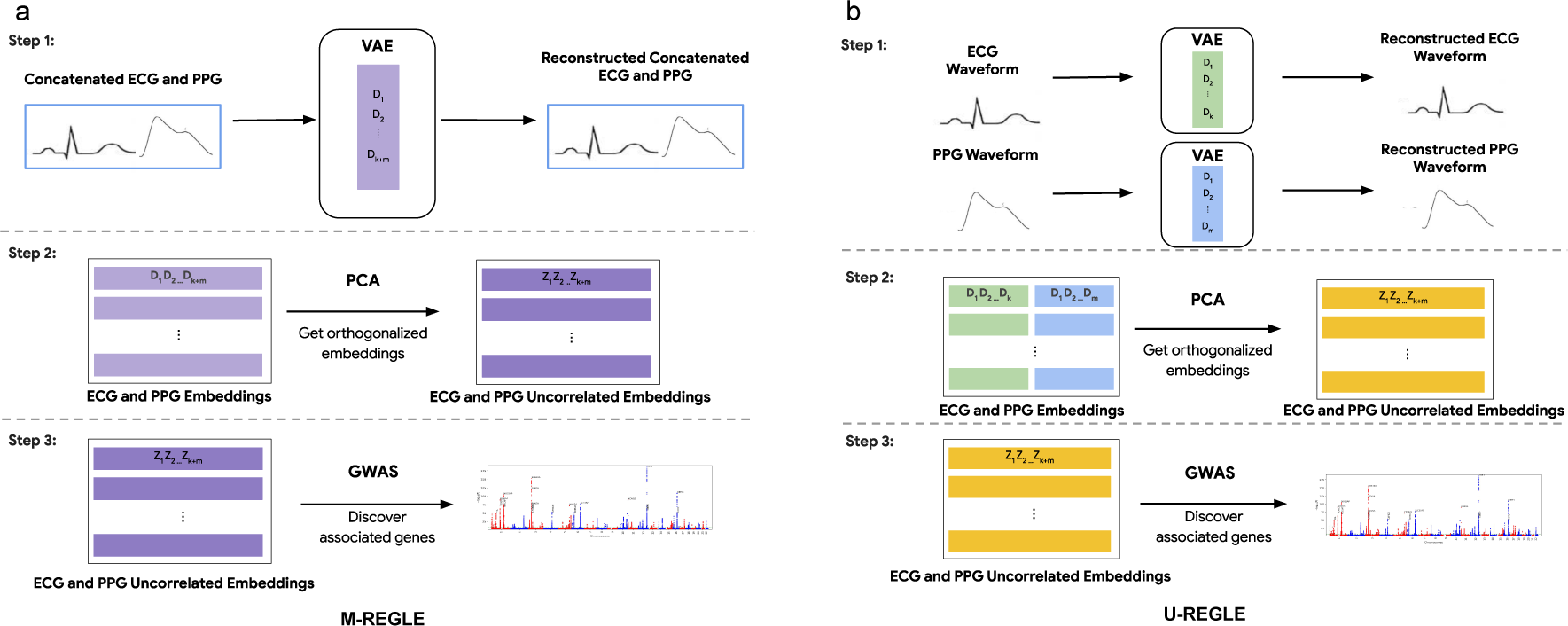
Overview of multimodal representation learning for genetic discovery on low-dimensional embeddings (M-REGLE). a) M-REGLE steps b) U-REGLE steps. Step 1 in M-REGLE (a) obtains the raw embeddings from multimodal HDCD in a joint fit while step 1 in U-REGLE (b) obtains the raw embeddings for each modality separately. In step 2, to ensure completely uncorrelated embeddings, we applied PCA on the raw embeddings. Lastly, we ran GWAS on the uncorrelated embeddings and combined them.

In the first step, we combined the multimodal HDCD (e.g., by concatenation), and used the combined data as input to learn a low-dimensional representation by using a VAE. The VAE model contains two parts: an encoder which encodes the input data into lower dimensional representations and a decoder which reconstructs the original data from the representation (Section 4). During training, the VAE model learned to compress the information from multimodal data into a latent embedding vector with a prior for uncorrelated coordinates that encouraged the VAE to learn disentangled embeddings. We obtained an *almost* disentangled joint representation for the input multimodal data. We observed a phenotypic correlation of *<* 0.13 for 12-lead ECG (Supplementary Figure 1) and *<* 0.05 for ECG Lead I + PPG (Supplementary Figure 2). In the second step, to ensure completely uncorrelated embeddings, we ran Principal Component Analysis (PCA) on the VAE embeddings to project them into completely uncorrelated PCs. In the final step, the PCs of the multimodal joint embeddings were used as synthetic phenotypes for GWAS. We performed GWAS on each PC of the embeddings for all individuals. We then combined the preliminary GWAS results by summing the chi-square statistics of each phenotype, deriving combined P-values, and forming combined hits and loci (Figure 1a). Finally, to obtain a polygenic risk score (PRS), we used elastic net regression to learn a linear combination of the hits. The main difference between M-REGLE and U-REGLE is that M-REGLE learns the embeddings of all modalities in a joint model while U-REGLE learns the embedding for each modality separately (Figure 1b). In addition to comparing M-REGLE with U-REGLE, we also compared the representations learned by multimodal and unimodal PCA and regular (non-variational) convolutional autoencoders (CAE).

### 2.2 Overview of cardiovascular modalities

We used the ECG and PPG HDCD in UK Biobank (UKB) to define two multimodal tasks. First, we used the at rest, median complex ECG waveforms from UKB. This median complex is the composite of several consecutive complexes that are aligned and of which the median value was taken for each timepoint of the cardiac cycle. Second, we extracted median heartbeat PPG waveforms from UKB. We studied two multimodal tasks: 1) 12 lead-ECG at rest, where the leads form 12 different modalities and 2) ECG lead I plus PPG (which are available in modern smartwatches). After performing quality control on the data (Section 4), we created train, validation, and test splits for model training, hyperparameter tuning and model evaluation, and PRS model evaluation, respectively (Supplementary Figures 3 and 4). Overall, the training, validation, and test datasets contain 70%, 20%, and 10% of samples and each dataset has similar phenotypic distributions (Supplementary Figure 5 and Supplementary Table 1).

Multimodal training is more effective in the presence of complementary information across modalities. To understand signal overlap and whether jointly learning embeddings is helpful, we performed the canonical-correlation analysis (CCA) to identify and measure the associations between the modalities. This measure ranges between 0 for no overlap and 1 for complete overlap. First, we performed both CCA and DCCA to compute the maximum projected correlation between each pair of the 12 leads of ECG. We observed that while some pairs have extremely high mapped correlation (e.g., 0.98 *±* 0.01 correlation between contiguous leads V5 and V6), other leads have comparatively lower correlation and thus contain complementary information (e.g., 0.62 *±* 0.01 correlation between limb lead III and precordial lead V2). Next, we analyzed ECG lead I plus PPG and observed a modest correlation, indicative of complementary signals (0.43 *±* 0.01) (Supplementary Table 2). As a reference point, we observed that the spirogram data–a measure of lung function–in UKB [7, 8] has a lower projected correlation with lead I ECG (0.29 *±* 0.03) and PPG (0.37 *±* 0.03). It is worth noting that we do expect to see some non-zero correlation as HDCDs capture general health information of individuals such as age, sex, and BMI. Lastly, we observed similar pattern when we utilized CCA extension called deep CCA [27] (DCCA), which relaxes the linear assumption in CCA (Supplementary Figure 6 and Supplementary Table 2).

### 2.3 M-REGLE produces better learned representations

When modalities have complementary and overlapping information, multimodal learning is expected to use the latent coordinates more effectively and thus produce better representations. One way to quantify a better representation is to have a lower overall reconstruction error, given a fixed latent dimension. We verified this hypothesis by applying Step 1 of M-REGLE and U-REGLE (i.e., jointly or separately learn representations from multimodal data) to both 12-lead ECG and lead I ECG and PPG tasks, and compared the reconstruction results.

We observed that M-REGLE reduced reconstruction error compared to U-REGLE on the validation set of the 12-lead ECG task. For M-REGLE, all the modalities needed to be combined as one input. We stacked the 12-lead waveforms as 12 channels (i.e., similar to the three channels of a color image). Then we trained a VAE model with 96 latent dimensions on the training data to reconstruct the 12-lead waveforms all together. For U-REGLE, we trained a VAE model independently on each ECG lead, getting 12 different VAE models. Each VAE had 8 latent dimensions. In total each sample had 12 *×* 8 = 96 latent dimensions, the same number as M-REGLE. We compared the MSE of the reconstructed 12-leads ECG by M-REGLE with the average MSE of the 12 ECG leads reconstructed by U-REGLE (which makes them comparable). M-REGLE reduced the overall MSE by 72.5%. The lower MSEs of M-REGLE were achieved consistently against different random seeds that were used in model training (Supplementary Figure 7a). We observed the same trend for different number of latent dimensions (Figure 2a and Supplementary Table 4). In addition, we compared multimodal PCA and CAE with unimodal PCA and CAE and the same trend was observed (Supplementary Figure 8 and Supplementary Tables 5 and 6).

**Figure 2:**
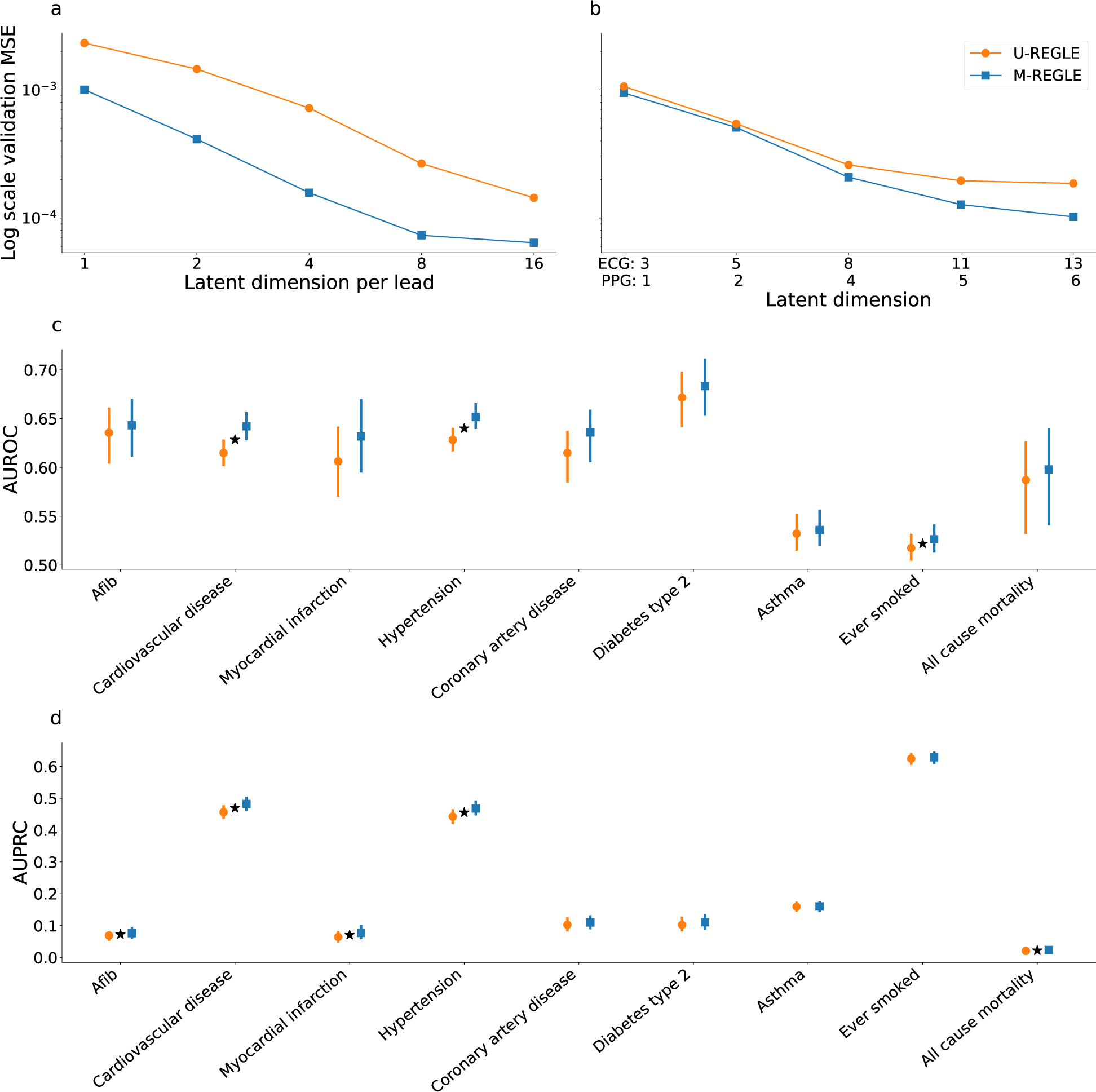
M-REGLE embeddings improve cardiovascular trait prediction. a) Validation reconstruction losses in log-scale of U-REGLE and M-REGLE on 12-lead ECG data. X-axis is the numbers of latent dimensions (1, 2, 4, 8, 16) per ECG lead. Standard errors (SE) are too small to plot (See Supplementary Table 4 for the SE). b) Validation reconstruction losses in log-scale of M-REGLE and U-REGLE across numbers of latent dimensions on Lead I ECG and PPG data. X-axis is the numbers of latent dimensions where the the first number is the latent dimension of ECG lead I and second number is latent dimension for PPG: 3+1, 5+2, 8+4, 11+5, 13+6 (See Supplementary Table 7 for SE of M-REGLE and U-REGLE reconstruction losses). All the difference between M-REGLE and U-REGLE in panels a and b are significant. c) AUCROC prediction of 9 phenotypes utilizing ElasticNet trained on the 12 embeddings obtained from ECG lead I and PPG, and d) AUPRC prediction of 9 phenotypes utilizing ElasticNet trained on the 12 embeddings obtained from ECG lead I and PPG. Star (*) sign indicates a statistically significant difference between the two methods using paired bootstrapping (100 repetitions) with 95% confidence.

The hypothesis was also verified on the lead I ECG and PPG task. We applied M-REGLE on ECG Lead I and PPG. To combine the two modalities, we concatenated ECG and PPG waveforms to form a long one channel input. We then trained a VAE model which learns to reconstruct the concatenated ECG and PPG waveforms. The VAE model generated 12 latent dimensions from the concatenated ECG and PPG waveforms (Section 4). In U-REGLE, we trained a VAE model for each modality independently. We used 8 latent dimensions in the ECG VAE model and 4 latent dimensions in the PPG VAE model. We assigned more dimensions to ECG, since ECG is more complex (Supplementary Table 3), and we observed comparable variance explained ratios in PCA when using 8 and 4 as PC numbers for ECG and PPG. We compared the overall reconstruction loss of M-REGLE with the weighted average of the U-REGLE reconstruction losses (weighted by the waveform lengths; Methods). M-REGLE achieved 20.23% lower overall reconstruction loss compared to U-REGLE, and this performance was robust against random seeds that models were trained on (Supplementary Figure 7b). We observed the same trend for different numbers of latent dimensions in VAE and different methods such as PCA and CAE (Figure 2b, Supplementary Tables 7 to 9 and Supplementary Figure 8).

### 2.4 M-REGLE embeddings improve cardiovascular traits prediction

To understand the M-REGLE embeddings, we computed the phenotypic correlation between MREGLE embeddings obtained from the 12 lead ECG and lead I plus PPG with 10,602 UKB phenotypes. Since these embeddings are highly associated with covariates age, sex, height, body mass index, and smoking status, we first removed (residualized) the effect of these covariates from MREGLE embeddings prior to computing the phenotypic correlation. We observed that 1,989 out of 10,602*108 M-REGLE embeddings and phenotype pairs had significant correlations after Bonferroni correction with very strong phenotypic correlations between M-REGLE embeddings and cardiac phenotypes indicating M-REGLE embeddings captured considerable cardiovascular information. Examples of these phenotypes were Position of the pulse wave peak (R=-0.69; P<1.6E-300) with 2nd embedding of PPG and ECG lead I, ventricular rate (R=0.50; P<1.77E-232) with 7th embedding of PPG and ECG lead I, R axis (R=-0.50; P<3.25E-232) with 96th embedding of 12-lead ECG, QT interval (R=-0.42; P<9.43E-158) with 4th embedding of PPG and ECG lead I, QRS num (R=0.37; P<3.80E-121) with 7th embedding of PPG and ECG lead I, Pulse rate (R=0.36; P<1.59E118) with 4th embedding of PPG and ECG lead I, heart rate during PWA (R=0.36; P<9.86E-117) with 7th embedding of PPG and ECG lead I (Supplementary Table 10).

To further investigate the correlation between M-REGLE embeddings and other phenotypes, we fit single-task Elastic Net models on M-REGLE embeddings to predict eight cardiovascular disease phenotypes and one trait ever smoked, which is known to be a main cardiovascular risk factor, in UK Biobank. We used the 12 M-REGLE embeddings obtained from lead I ECG and PPG. We observed that in the case of the lead I plus PPG embeddings, for all 9 phenotypes, M-REGLE outperformed U-REGLE in terms of AUROC and AUPRC. (Figure 2c, 2d and Supplementary Tables 11 to 14). Based on a paired bootstrap analysis, M-REGLE was significantly better than U-REGLE, in terms of AUROC, for predicting hypertension, cardiovascular disease, and ever smoking, while M-REGLE was significantly better than U-REGLE, in terms of AUPRC, for predicting for afib, cardiovascular disease, myocardial infarction, hypertension, and all cause mortality.

### 2.5 M-REGLE on 12-lead ECG enhances genetic discovery

We compared the genetic discovery performance of M-REGLE with U-REGLE on the combined training and validation set (n = 38,697). M-REGLE generateed 96 dimensional representations for each individual and then PCA was performed to project the raw embeddings into 96 uncorrelated PCs (i.e., there was no loss of information and this was only for orthogonalization). We performed GWAS on each of the 96 PCs using REGENIE [28] (Supplementary Table 15). As the 96 PCs were uncorrelated, we combined these 96 GWAS by summing the chi-square statistics and computed the final combined p-value from this summation by choosing 96 degrees of freedom [29]. We used the combined p-values to create the hits and loci (Methods). Hits were independent genome-wide significant (GWS) variants (*R*^2^ *≤* 0.1 and *P ≤* 5 *×* 10*^−^*^8^) and loci were obtained by merging hits within 250 kb together (Figure 3a).

**Figure 3:**
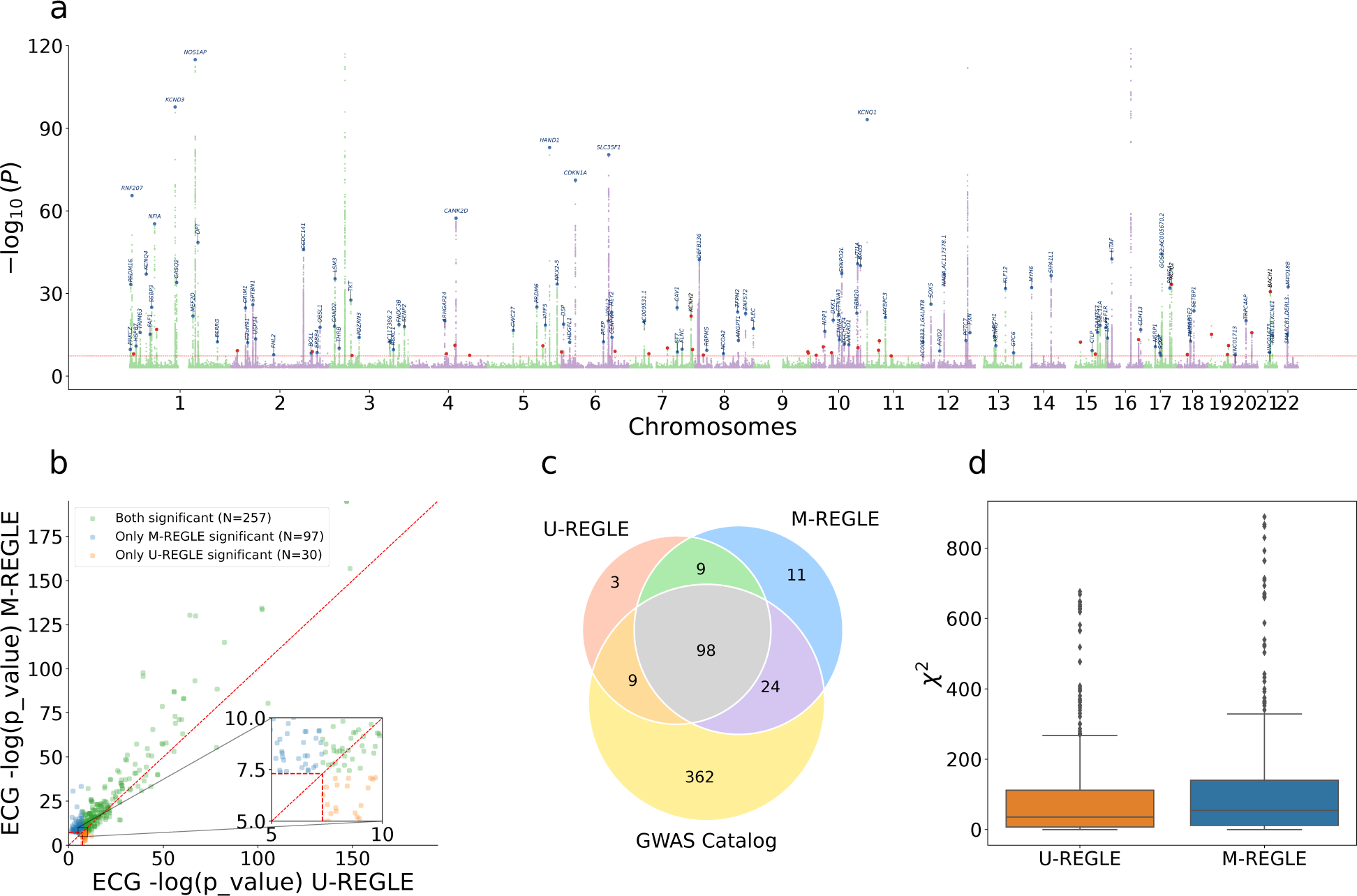
M-REGLE on 12 ECG leads increases genomic discovery. a) Manhattan plot depicting M-REGLE GWAS p-values for all 22 autosomal chromosomes. Black gene names indicate the closest gene for each locus with *−* log_10_ *p >* 20 and red dots denote all other GWS loci. Blue gene names and dots indicate loci also identified in U-REGLE. b) Comparison of M-REGLE GWS variants-in-hits with U-REGLE. The X-axis is the *−* log p-value of U-REGLE. The Y-axis is the *−* log p-value of the M-REGLE. All p-values in (a) and (b) are computed by summing the chi-square statistics for all 96 embeddings to perform a single joint chi-square test. The vertical and horizontal red lines indicate the GWS level. The diagonal red line indicates *y* = *x*. The orange dots indicate variants-in-hits that are significant for U-REGLE but not significant for our M-REGLE and green dots indicate variants-in-hits that are significant for our M-REGLE but not significant for U-REGLE. c) A 3 way Venn diagram of the GWAS catalog loci, loci discovered by M-REGLE and loci discovered by U-REGLE. d) Comparison of the chi-square statistics for all known significant variants in GWAS catalog for both U-REGLE and M-REGLE. The difference is statistically significant.

M-REGLE had higher strength of association compared to U-REGLE (Figure 3b and Supplementary Table 16) where M-REGLE discovered 262 hits (62 more than U-REGLE) and 142 loci (23 more than U-REGLE) (Figure 3c and Supplementary Table 17). To compare our results with known loci previously detected for ECG and cardiovascular traits (Supplementary Table 33), we compiled all the ECG-related trait hits (n=989) and loci (n=493) from the GWAS Catalog (Section 4). From 262 hits and 142 loci discovered by M-REGLE, 231 (88.5%) hits and 122 loci (85.92%) were previously reported (Figure 3c and Supplementary Table 18) leaving 20 loci genome-wide not previously associated with these traits. To compare GWAS statistical power, we calculated the expected chi-square statistics on all the variants reported as ECG-related traits in GWAS catalog. We observed that MREGLE had higher statistical power than U-REGLE (*E*[*χ*^2^] = 112.48 *±* 2.05 vs *E*[*χ*^2^] = 92.2 *±* 1.83) (Figure 3d and Supplementary Table 17).

As a secondary analysis, we used PCA and CAE in multimodal and unimodal settings to obtain latent representations. We observed similar results where multimodal learning leaded to better genetics discovery and higher statistical power. Multimodal PCA discovered 215 hits and 122 loci, while unimodal PCA found 167 hits and 104 loci (Supplementary Table 17). Similarly, most of the hits and loci were known to be ECG-related traits (Methods). As for statistical power, the expected chi-square statistic of Multimodal PCA was 90.98 *±*1.70, which is significantly higher than that of unimodal PCA (*E*[*χ*^2^]=71.53 *±*1.38). We observed similar trends for CAE (Supplementary Table 18).

To assess functional enrichments, we used GREAT [30] to quantify the number of cardiovascular terms significantly associated with loci identified by each method as well as the statistical significance of enriched terms. For both multimodal and unimodal models, REGLE outperformed the corresponding PCA method in all cases, and outperformed CAE in 3 of the 4 cases with respect to cardiovascular term enrichments (Supplementary Table 19) and overall term significance (Supplementary Table 20).

### 2.6 M-REGLE on ECG lead I and PPG enhances genetic discovery

Similar to the 12 lead ECG case, in the ECG lead I and PPG task, we observed that M-REGLE had stronger genetic discovery results than U-REGLE. We generated 12 dimensional joint representations for all individuals in the training and validation set (n = 33,192) by running the M-REGLE model on concatenated ECG lead I and PPG waveforms, then performed PCA to obtain 12 uncorrelated PCs of the joint representations. We performed GWAS using REGENIE on each of the 12 PCs (Supplementary Table 21) and combined the GWAS result by summing the chi-square statistics of the 12 PC GWAS (Figure 4a).

**Figure 4:**
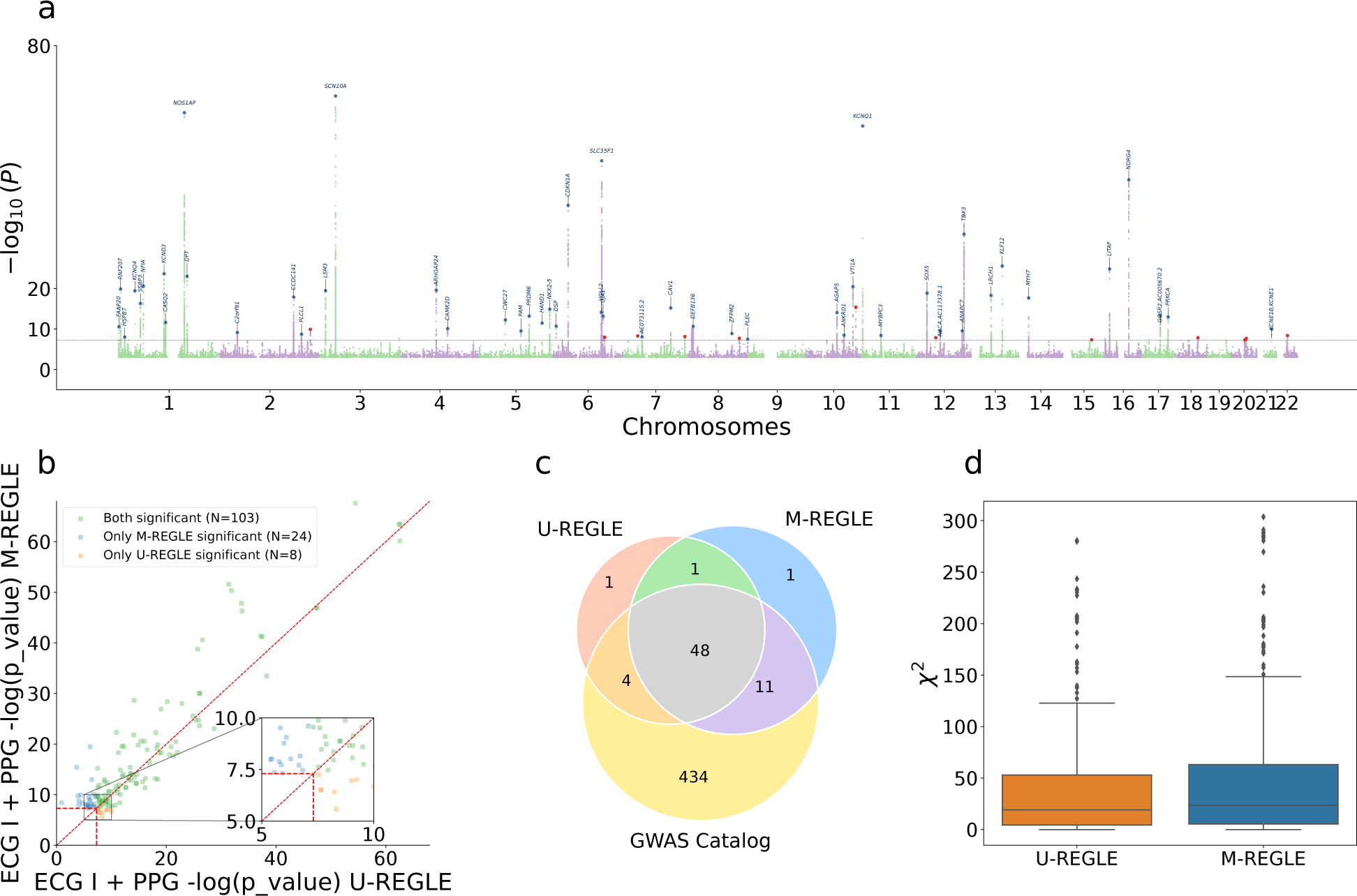
M-REGLE on ECG lead I and PPG increases genomic discovery. a) Manhattan plot depicting M-REGLE GWAS p-values. Black gene names indicate the closest gene for each locus with *−* log_10_ *p >* 20 and red dots denote all other GWS loci. Blue gene names and dots indicate loci also identified in U-REGLE. b) Comparison of M-REGLE GWS variants-in-hits with U-REGLE. The X-axis is the *−* log p-value of Baseline. The Y-axis is the *−* log p-value of the M-REGLE. All p-values (a) and (b) are computed by summing the chi-square statistics for all 12 embeddings to perform a single joint chi-square test. The vertical and horizontal red lines indicate the GWS level. The diagonal red line indicates *y* = *x*. The orange dots indicate variants-in-hits that are significant for U-REGLE but not significant for our M-REGLE and green dots indicate variants-in-hits that are significant for our M-REGLE but not significant for Baseline. c) A 3 way Venn diagram of the GWAS catalog loci, loci discovered by M-REGLE and loci discovered by U-REGLE. d) Comparison of the chi-square statistics for all known significant variants in GWAS catalog for both U-REGLE and M-REGLE. The difference is statistically significant.

**Figure 5:**
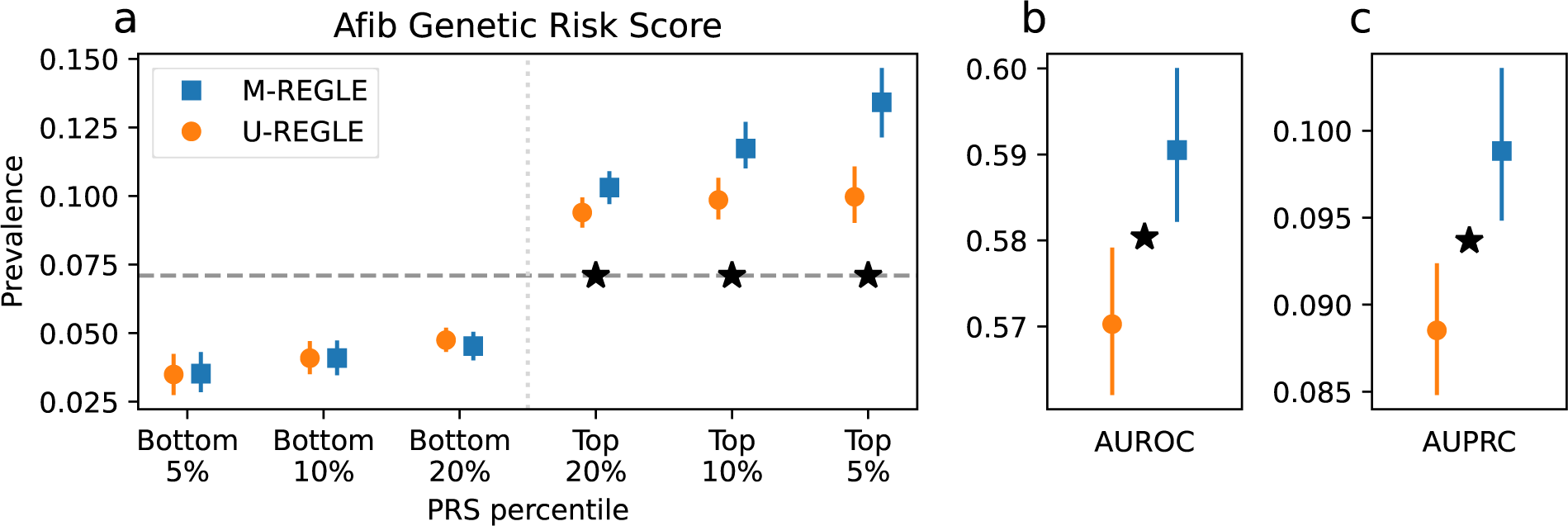
M-REGLE improves Afib genetic risk score. a) X-axis is genetic risk score percentile and Y-axis is the prevalence. Lower is better for the bottom percentiles; higher is better for the top percentiles. b) AUROC, and c) AUPRC (precision recall) Star (*) sign indicates a statistically significant difference between the two methods using paired bootstrapping (100 repetitions) with 95% confidence.

M-REGLE has higher strength of association compared to U-REGLE (Figure 4b) where GWAS on M-REGLE embeddings detected 103 GWS hits and 61 loci, 14 and 7 more than U-REGLE, respectively (Supplementary Table 22). We compared the loci with GWAS Catalog. Out of 61 GWS loci detected by M-REGLE from lead I ECG and PPG, 59 fall in the known loci (96.72%), and 2 were not previously known (Figure 4c and Supplementary Table 23). To further show that M-REGLE improves statistical power, we calculated the expected chi-square statistics on all GWAS catalog variants for ECG and PPG related traits. We observed a significant power increase for M-REGLE (*E*[*χ*^2^] = 51.66 *±* 0.98) compared to U-REGLE (*E*[*χ*^2^] = 44.37 *±* 0.88) (Figure 4d and Supplementary Table 24). We observed the same pattern for CAE, but multimodal and unimodal PCA were comparable (Supplementary Table 24).

Similar to 12-lead ECG, functional enrichments of cardiovascular terms for ECG lead I and PPG models showed superiority of multimodal and unimodal REGLE over PCA and CAE modeling methods for both cardiovascular term enrichments (Supplementary Table 19) and overall term significance (Supplementary Table 20).

### 2.7 M-REGLE improves the PRS for cardiovascular traits

To ensure the novel hits detected by M-REGLE are biologically important, we created genetic risk scores by extracting all variants from the significant hits. For each set of embeddings, we trained an ElasticNet model to predict the phenotype of interest on the train dataset utilizing all significant variants (see Methods). We considered multiple ECG-derived (e.g., ECG QT interval, ECG P axis, etc) and PPG-derived (e.g., PPG pulse velocity) phenotypes as well as 7 additional cardiovascular disease or cardiovascular risk phenotypes for which there is extensive knowledge available on genetic associations such as atrial fibrillation (Afib), BMI, height, hypertension, stroke, T2D, Systolic Blood Pressure (SBP). In UKB, we have 22 derived phenotypes from ECG and PPG while in total we utilized 29 phenotypes.

We observed that genetic risk for 5/29 phenotypes were significantly improved by M-REGLE compared to U-REGLE after Bonferroni correction (Supplementary Table 25). The most notable improvement was observed for Afib, with M-REGLE showing an AUROC of 0.58 (95% CI: 0.582-0.599) versus U-REGLE AUROC of 0.56 (95% CI: 0.562-0.578) and an AUPRC of 0.10 (95% CI: 0.094-0.103) versus 0.09 (95% CI: 0.084-0.092) when utilizing all 12 ECG leads (Supplementary Table 25). Additionally, other phenotypes such as PPG pulse rate, ECG QT interval, SBP, and height also exhibit significant improvement in phenotypic prediction with M-REGLE loci compared to U-REGLE loci (Supplementary Table 25).

To further validate our PRS results, we computed the PRS in Indiana Biobank datasets [31], European Prospective Investigation into Cancer (EPIC)-Norfolk [32], and the British women’s heart and health study (BWHHS) [33–35] using the weights learned in the UK Biobank (Methods). In the Indiana Biobank [31], we have access to Afib and observed that for all metrics M-REGLE outperforms U-REGLE among European samples. In the case of AUROC, M-REGLE was significantly better (Supplementary Table 26). In EPIC-Norfolk [32], we have access to Afib, PPG pulse rate, ECG QT interval, and SBP. We observed that M-REGLE outperformed U-REGLE for all metrics. For Afib in particular, the AUROC, AUPRC, and Top 5% prevalence were significantly better for M-REGLE compared to U-REGLE (Supplementary Tables 27 and 28). Similar to EPIC-Norfolk, in BWHHS, we have access to Afib, PPG pulse rate, ECG QT interval, and SBP. We observed that for all metrics M-REGLE outperforms U-REGLE (Supplementary Table 29 and for Afib AUPRC and Top 5% prevalence were significantly better for M-REGLE compared to U-REGLE (Supplementary Table 29). Lastly, for all other phenotypes M-REGLE outperformed U-REGLE except for SBP in BWHHS (Supplementary Table 30).

## 3 Discussion

Access to multiple high dimensional clinical data (HDCD) modalities for each individual in wearable biosensors, biobank, and EHR systems provides unique opportunities to study the genetics of complex traits. In particular, when several data modalities are related to a single organ system, these modalities together can provide a better picture of the organ’s function than any individual modality. However, we currently lack statistical methods to fully utilize these multimodal HDCD in genetic analyses. REGLE (Yun et al.) [8] provides a means to study the genetic underpinnings of high dimensional data, but it is limited to one data modality, and does not leverage the shared and complementary information in multimodal HDCD. Here, we developed an unsupervised representation learning method, M-REGLE, which aims to utilize multimodal HDCD in a joint model to improve genetic analyses. We showcased the advantages of M-REGLE over applying REGLE independently on different HDCD modalities and then combining the results.

We compared M-REGLE with the unimodal representation learning analog (U-REGLE) side-by-side on the same input data and we observed significant performance improvements in several aspects. First, M-REGLE has lower reconstruction error compared to U-REGLE, which suggests it compresses and keeps more of the available signal in the dense representations for subsequent use in GWAS. Second, we validated that the joint embeddings learned by M-REGLE capture more genetic signals related to the input data compared to the collective U-REGLE embeddings (Figures 3d, 3c, 4d, and 4c) and higher statistical power over known cardiovascular traits (Figures 3d and Figure4d). Lastly, the genetic variants identified by M-REGLE enable better polygenic scoring for 14% (4/28; excluding height from 5/29 phenotypes reported in the Result section) of tested cardiac phenotypes. Overall, M-REGLE provided greater statistical power for GWAS on HDCD, which could improve downstream applications such as biological discovery, better PRSs, and more possible drug targets

The key for the effectiveness of M-REGLE is the existence of complementary and overlapping information in the modalities. On one end of the spectrum, when the information in different modalities is completely non-overlapping, there is no duplication of information (and consequently inefficiency). Hence, learning separately will not be wasteful and M-REGLE is not expected to learn more effectively than U-REGLE. On the other hand, when some modality completely covers another modality, we will not expect to gain any boost from using M-REGLE either, because the duplicated information does not add a new signal to the learned representation. HDCDs of a single organ system or disease are usually measured through different methods and sensors, and fall in the middle of this spectrum. However, before applying M-REGLE, we recommend performing a preliminary analysis to check that the modalities include complementary information(e.g. by performing CCA or DCCA).

With the presence of complementary and overlapping information, we expect that when using a deep neural network, the compression of information in a low-dimensional dense representation happens more effectively and as a result, we retain more signal that translates into better reconstruction by the decoder. Thus, the key question was whether the more effective compression also translates into better genetic discovery. We hypothesized the answer would be positive, because by using more than one source, the signal to noise ratio can be improved, which translates into better statistical power. On the one hand, the representation learning will be more efficient and can retain more information that improves the signal. Furthermore, the noise can be reduced because one modality can correct the noise in the other. Our results using different data modalities, different learning methods, and multiple biobanks corroborate this hypothesis.

It is worth noting that our work has a connection to fusion models in deep learning. When dealing with multimodal data, there are three common paradigms often used when combining modalities. First, in early fusion, one combines modalities before the representation learning process and learns a joint latent that is used for the final task, such as prediction of Afib status. Second, in intermediate fusion, one learns representations for each modality separately, combines the independent representations (e.g., by concatenation), and performs the final task on this combined representation. Third, in late fusion, one both learns the representations separately and also uses the separate latents to obtain a separate result using each modality for the final task. One then combines the final results. For example, we use PPG and ECG separately to obtain two separate probabilities for having Afib and then get their average as the final probability. Note that M-REGLE is an early fusion, while U-REGLE is an intermediate fusion. In contrast, a late fusion method would be to run original REGLE on each modality separately (which includes the final task of GWAS), and then meta-analyze the resulting separate GWASs. We intentionally choose U-REGLE (i.e. intermediate fusion) as the baseline instead of the late fusion of separate REGLEs. This is to limit the difference to only multimodal vs unimodal learning and have a better comparison.

Our exploratory work on ECG and PPG data also opens up opportunities in the large-scale study of cardiovascular disease risk and the genetics of phenotypes derived from smart wearable or mobile phones. Most of the popular smart wearable devices contain sensors which allow users to easily record their ECG (equivalent to the lead I ECG) and PPG. Our experiments demonstrate that M-REGLE will enable researchers to optimally leverage this newly abundant data for genetic discovery. In particular, we emphasize the importance of early fusion of the data before the representation learning and compression into low-dimensional data as there is a large gap between before and after fusion. This is in contrast with usual practice of using meta-analysis after GWAS to combine multiple sources of data in genetic analysis.

Our work has several limitations. First, our aim was to focus on the method and illustrate the advantage of M-REGLE for genetic discovery over combining the results of unimodal representation learning. As a result, many aspects of genetic discovery can be improved and the goal was not to have the strongest GWAS on cardiovascular phenotypes. Second, we utilized the median of waveforms of heartbeats for both PPG and ECG. Alternative preprocessing methods can be explored on full ECG and PPG waveforms, which potentially could produce more comprehensive representations, when the focus is on the genetic architecture of heart function. Third, we performed the main experiments on samples with all the modalities available and have not fully explored M-REGLE’s potential in handling missing data. Being able to learn joint representations from a multimodal dataset with missing data would enable us to carry out larger scale genetic studies which cover all individuals with only some of the data modalities available. Fourth, we observed that the value of *β* in *β*-VAE and scaling factor of inputs jointly have significant impact on M-REGLE embeddings (Supplementary Figures 9 and 10). While we used a fixed Scaling factor, and performed hyperparameter search over *β* to find the optimal *β* for the given scaling factor. A more systematic search over both *β* and scaling factor can produce better embeddings.

Despite these limitations, M-REGLE provides a means to better leverage multimodal HDCD for genetic discovery, and has shown improved performance compared to unimodal representation learning. We demonstrated its advantages by using ECG and PPG data to study genetics of cardiovascular traits. We believe M-REGLE will become a standard method for using multimodal HDCD for GWAS and downstream analyses.

## 4 Methods

### 4.1 Data Preparation

12-lead electrocardiogram (ECG) data were sourced from UKB field 20205. We used the at rest, median heartbeat (median complex) waveforms. This median complex is the composite of several consecutive complexes that are aligned, and of which the median value was taken for each time-point of the cardiac cycle. Each median waveform contains 600 points for all leads. The median data are relatively cleaner compared to the raw ECG full waveform; therefore, it did not require extensive preprocessing. The photoplethysmogram (PPG) waveform was extracted from UKB field 4205, but unlike ECG, the PPG data have only the median heartbeat waveform. Each PPG waveform contains 100 points, which are median values from single heartbeats. We made two multimodal datasets from the data: 1) 12 leads ECG as 12 different modalities, 2) lead I of ECG and PPG. For both 12-lead ECG and PPG, we used data Instance 2 in UKB, which contains the most overlapping samples. We obtained 50,469 samples for 12-lead ECG and 55,041 samples of PPG as of April 17, 2023.

We first applied Finite Impulse Response Filters (FIR) on all the ECG data to reduce noise. FIR filters are widely used in signal processing for removing unwanted frequency components from a signal. We set the cutoff frequencies 0.05 and 40. We only performed this step on ECG, because it contains more noise compared to PPG. For quality control, we calculated several statistics (minimum, maximum, mean and median) on each waveform, and compared them with the statistics of all the other waveforms of the same type. We set a cutoff percentile of 0.1 and 99.9 for each statistic. The waveforms with at least one statistic that fell outside of these cutoffs were dropped. After this step, we had 48,259 samples for the 12-lead ECG and 41,282 samples in the ECG lead I and PPG dataset.

### 4.2 Dataset generation for machine learning model

For both datasets, we created train/validation/test splits for hyperparameter tuning, model evaluation and a final test for the PRS model. We first created the splits by using a static hash on all the available EIDs. Each EID was assigned to one of the training, validation and test sets. Since we expect a uniform distribution from a hash function, the splits should exhibit the same properties for any feature of interest (e.g. the age of the individuals). The datasets were split by looking up each of the EIDs in the datasets in the created splits. The training, validation and test dataset contained 70%, 20% and 10% of the samples, respectively.

In order to standardize the magnitudes of waveforms from different modalities, we proceeded to scale each waveform type, ensuring their values fell approximately within [*−*1, 1]. We calculated the scale factor of each modality separately, by picking the 90th percentile of the absolute maximum values of all waveforms of the modality in the training dataset. Therefore, we avoided the impact of outlier waveforms with extreme maximum values. We then applied the scales for each modality on all training, validation and test datasets, by dividing the waveforms with the corresponding scale factor.

### 4.3 Learning 12-lead ECG representations with generative models

Variational autoencoder (VAE) models were trained to learn lower-dimensional representations from 12-lead ECG data. Autoencoders comprise as encoder and decoder function approximators linked by a narrow bottleneck layer, autoencoders condense input data into a concise set of numbers at the bottleneck layer, with the decoder subsequently reconstructing the input data from this condensed representation [36]. VAEs [37] represent a distinctive variant of autoencoders, introducing stochasticity into the encoding process. For M-REGLE, one VAE model was trained on the combined 12-lead ECG data to reconstruct the 12 leads together. For U-REGLE, 12 separate VAE models were trained on each ECG lead.

We used similar model structures for M-REGLE and U-REGLE. The encoder of VAE models consists of 1D convolutional layers, each followed by max pooling, then fully-connected layers to generate the mean and variance of the bottleneck layer. The decoder architecture is a mirror image of the encoder. It starts with fully-connected layers, followed by transpose convolutional layers, each prepended by an upsampling layer. For unimodal VAEs, the ECG lead was directly used as one input channel. In M-REGLE, the 12 leads were first stacked to create an input in the shape of 600 points x 12 channels. The VAE models for single lead ECG all had 8 latent dimensions. We explored the latent dimension number by performing PCA using various PC numbers on the ECG leads. When we used 8 PCs, it explained 0.93 of total variance of the data, demonstrating that 8 latent dimensions is sufficient for a linear model to learn good representations from single ECG leads. We used 96 latent dimensions for multimodal VAE, so that the total number of dimensions in the unimodal representations and multimodal representations were the same. The standard VAE loss function consisting of the reconstruction loss and the (rescaled) Kullback–Leibler (KL) divergence loss were used as the objective during the training of all the VAE models. The Adam optimizer was used to optimize the objective.

For each of the VAE models, we performed a large-scaled hyper-parameter search independently (Supplementary Table 31). Each unimodal VAE model was trained for at most 300 epochs. The multimodal VAE models were trained for at most 1200 epochs. After training, the checkpoints with the lowest validation loss were selected. Because one advantage of the VAE model is its ability to learn disentangled representations, we also used how uncorrelated the resulting representation coordinates were as a metric to pick the hyper-parameter sets. We calculated the correlation matrix of the representations that the model checkpoint generates on the validation dataset, and found the maximum absolute value of the correlation coefficients (only among the non-diagonal values of the correlation matrix), which we use as the metrics for the disentangleness of generated representations. To decide which hyper-parameter set to use, we first set a threshold of 0.1 on the maximum absolute correlation value, and chose the one with the lowest validation reconstruction loss among all the models that passed the maximum absolute correlation threshold (Supplementary Table 32). To ensure our models were robust against random seeds, we retrained the selected hyper-parameter combination of every modality and data setting on 100 different random seeds. The retrained models all reached similar reconstruction loss and overall loss on the validation dataset (Supplementary Figure 7a).

As a baseline, PCA were trained on each of ECG 12-lead separately (unimodal PCA), and a combination of ECG 12-lead (multimodal PCA). We used 8 PC for PCA on single ECG lead, and 96 PC numbers for the PCA on joint ECG 12-lead. The learned PCs were used in the same way as the lower representations learned by VAE models in GWAS and downstream analyses.

### 4.4 Learning lead I ECG and PPG representations with generative models

VAE models were trained to learn representations of ECG lead I and PPG. We compared M-REGLE, where we trained one VAE model to reconstruct both waveforms together, and U-REGLE, where we trained two separate VAE models to learn representations of the waveforms independently.

We used the same model structure here as the 12-lead ECG models, which consist of 1D convolution layers and fully connected layers. For unimodal VAEs, the waveform was directly used as a one-channel input. In M-REGLE, we connected each individual’s PPG waveform to their ECG waveform to form a 700 points long one channel input. We used 8 latent dimensions in the unimodal ECG model, 4 latent dimensions in the unimodal PPG model, and 12 latent dimensions in M-REGLE. We selected the latent dimension numbers by experimenting with different PC numbers (Supplementary Table 3).

The training, model selecting strategy and the baseline creation of ECG lead I and PPG followed the same procedure as the 12-lead ECG (Supplementary Table 31, Supplementary Table 32, Supplementary Figure 7b).

Similar to the 12-lead ECG setting, we also created baselines using multimodal PCA and unimodal PCA. For multimodal learning, we first concatenated the ECG Lead I and PPG waveform, then learned a PCA transformation which reduces the data to 12 dimensions. For unimodal learning, we learned a PCA which generates 8 PCs for ECG Lead I and a PCA which generates 4 PCs for PPG.

### 4.5 Genome-wide association studies

We generated unimodal representations and multimodal representations for the 12-lead ECG and the ECG lead I + PPG on the combination of the training and validation datasets. We performed PCA on each set of representations of the dataset to transform the representations as uncorrelated coordinates. Then preliminary GWASes were performed on each of the PCs of the representations for each data setting and modality type using REGENIE [28]. The GWASes were adjusted for age, sex, body mass index, smoking status, genotyping array and the top 15 genetic PCs [7, 8].

To get an overall result of genetic discovery on the multimodal waveform data, we combined the preliminary GWASes on the PCs of the representations. We computed the summation of the chi-square statistics for each GWAS and computed the combined p-value by applying the survival function of the combined chi-square statistics, using the number of the phenotypes as the degree of freedom. We utilized the combined chi-square statistics to obtain the final GWAS result. More specifically, we considered that we had access to *m* phenotypes denoted by *P* = [*P*_1_*, P*_2_*,…P_m_*] that were not correlated and where 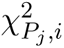 indicated the chi-square statistics of i-th SNP for *P_j_* phenotype. As phenotypes were not correlated, we computed the summation of all phenotypes chi-square statistics and computed the final p-value assuming that the summation followed a chi-square distribution with degree freedom of 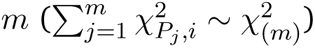 [29].

GWAS was restricted to individuals with European ancestry to minimize confounding. For quality control, we retained variants with minor allele frequency *≥* 0.001, imputation INFO score *≥* 0.8, missing call fraction *≤* 0.05, and Hardy-Weinberg equilibrium P>1.0E-10, among all genotyped and imputed variants provided by UK Biobank (UKB). Genome-wide significant “hits” were defined as the most significant variants with P<5.0E-8 and independent at *R*2 *<* 0.1 using the PLINK –clump command. A reference panel for linkage disequilibrium (LD) calculation contained 10,000 unrelated European samples from the UKB. Significant “loci” were created based on the span of reference panel SNPs in LD (R2 *≥* 0.1) with the hits. Loci separated by fewer than 250 kb were subsequently merged.

### 4.6 GWAS Catalog Loci Replication

The number of hits and loci may be misleading because some of them might be false-positives. However, if the hits/loci are known to be related to ECG or PPG traits, they are less likely to be false positive. We investigated relevant hits/loci by examining if they are previously reported in the literature to be ECG or PPG related. We performed a GWAS catalog search using key words (case insensitive), for example, “electrocardio”, “ecg”, “pr interval”, “ventricular rate”, “pulse wave”, “notch position”, etc (Supplementary Table 33).

### 4.7 GWAS Power Analysis via expected chi-square statistics

We compared the GWAS power of M-REGLE and U-REGLE in terms of the high-quality hits, by calculating the mean of the chi-square stats of all the rediscovered hits (i.e. the ones with support by GWAS catalog) from the GWAS of each modality and data setting.

### 4.8 Functional enrichment using GREAT

GREAT enrichment analyses were performed on the human GRCh37 assembly using GREAT v4.0.4 [30] using program defaults. Terms were considered statistically significant if the Bonferroni-corrected P-values for both the region-based and gene-based tests were *≤* 0.05. Cardiovascular terms were identified by performing a case-insensitive match on enriched ontology term descriptions using the regular expression “cardiac|cardio|cardial|heart|circulatory”. Paired T-tests were performed on *−log*_10_-transformed raw region-based P-values for statistically significant cardiovascular terms.

### 4.9 Creating polygenic risk scores from significant hits

To evaluate the improvement of one model over another (e.g., M-REGLE vs U-REGLE), we utilized phenotypic prediction as a measure of comparison. We created polygenic risk scores (PRSs) from the significant hits obtained for each model by training an ElasticNet model. The significant hits reported by each model were used as features for ElasticNet, with the prediction target set as the phenotype of interest (e.g., Afib). The same training set that was used to train M-REGLE was used to fit ElasticNet and we performed 5-fold cross-validation to find the right hyperparameters. We applied sklearn.linear_model.ElasticNet to train our ElasticNet and performed a hyperparameter search of l1_ratio over [0.1, 0.5, 0.7, 0.9, 0.95, 0.99, 1.0] values.

All the PRS values reported in the UK Biobank were obtained from the test dataset. Furthermore, we shared the variant weights derived from significant hits in the UK Biobank with our collaborators for assessment on the EPIC-Norfolk [32] and Indiana Biobank datasets [31]. Our collaborators assessed PRS across the phenotypic data available to them. Consequently, we obtained phenotype prediction results for the Afib phenotype in the Indiana Biobank dataset [31], while in the EPIC-Norfolk dataset, we obtained results for Afib, PPG pulse rate, and SBP (systolic blood pressure).

### 4.10 PRS validation on multiple datasets

#### Indiana Biobank dataset

Indiana Biobank samples were genotyped using Illumina Infinium Global Screening Array (GSA, Illumina, San Diego, CA) by Regeneron (Tarrytown, NY). Variants with palindromic alleles, missing rate >5%, minor allele frequency (MAF)<3%, and Hardy-Weinberg equilibrium P<1E-4 were excluded. Principal components (PC) of population stratification were calculated using Eigenstrat [38]. Based on the first two PCs, those clustered with the European reference samples from the 1000 Genomes Project [39] were grouped as EA samples and they were used in this study (N=4,030). Indiana Biobank samples were imputed using the Michigan Imputation Server [40] with the 1000 Genomes project as the reference panel. Variants with imputation INFO <0.3 or MAF <0.01 were excluded. PRS were calculated by using imputation dosages. Atrial fibrillation cases (N=778) were determined based ICD-9 (427.3, 427.31, and 427.32) and ICD-10 (I48.0, I48.1, I48.2, and I48.9) codes. Those not having atrial fibrillation in their electronic health records were considered as controls (N=3,252).

#### EPIC-Norfolk dataset

Genotyping in EPIC-Norfolk used the Affymetrix UK Biobank Axiom Array with the Axiom GT1 algorithm used for calling. Quality control exclusions were made for variants with Hardy–Weinberg P<1.0E-6 and checks made for abnormal clustering or plate batch effect. Samples were excluded for SNP call rate < 97% with further checks for heterozygosity and gender. Imputation was performed by the Sanger Imputation Service using the IMPUTE4 software and Haplotype Reference Consortium and UK10K plus 1000 Genomes phase 3 reference panels. Variants with a genotyping quality score (INFO) < 0.4 were excluded. Atrial fibrillation (N=3,554) was ascertained using Hospital Episode Statistics (HES) records maintained in a database containing medical records for all UK National Health Service (NHS) hospitals. Linkage to cohort participants used the unique NHS number. Cases were defined as ICD-10 I48.0, I48.1, I48.2, I48.9 or I48 unspecified (coded as I48X). Controls (n=18,002) were defined as non-cases with available GWAS. *British women’s heart and health study (BWHHS):* BWHHS utilized genotyping techniques employed in the Human cardiovascular disease (HumanCVD) BeadChip, also known as the ITMAT-Broad-CARe (IBC) v2 array. This array contained up to 49,240 SNPs. To ensure the reliability of the data, rigorous quality control measures were implemented. All SNPs were clustered into genotypes via Illumina Beadstudio software and stringent quality control filters were applied at both the sample and SNP level. Samples failing to meet criteria, including individual call rates below 90%, gender mismatches, and duplicate discordance, were excluded. Similarly, SNPs with call rates below 95% or exhibiting Hardy-Weinberg disequilibrium with a P < 1.0E–6 were removed. Furthermore, to capture low-frequency variants of significant effect across the extensive dataset, filtering was conducted based on a minor allele frequency (MAF) threshold of less than 0.01. Incidence cases of atrial fibrillation (AF) were identified through general practitioner (GP) information provided during BWHHS record reviews. Additionally, prevalent AF cases were identified through ECG diagnosis, ensuring a comprehensive assessment of AF prevalence within the study population.

## Code Availability

Open-source code and trained model weights are available at https://github.com/Google-Health/genomics-research under the mregle directory.

## Supporting information

Large Supplementary Tables

## Data Availability

Datasets used in this study (UK Biobank, EPIC Norfolk, Indiana Biobank, and British Women's Heart and Health Study) are available to qualified researchers via applying access to each dataset maintainers. Open-source code and trained model weights are available at https://github.com/Google-Health/
genomics-research under the mregle directory. M-REGLE values of UK Biobank individuals will be returned to UK Biobank and will be made available by UK Biobank https://www.ukbiobank.ac.uk/.

https://github.com/Google-Health/genomics-research

https://www.ukbiobank.ac.uk/

## Acknowledgements

We thank all participants, dataset creators, and maintainers of UK Biobank, BWHHS, EPIC Norfolk, and Indiana Biobank. This research has been conducted using the UK Biobank Resource under Application Number 65275. The EPIC-Norfolk study (DOI 10.22025/2019.10.105.00004) has received funding from the Medical Research Council (MR/N003284/1 MC-UU_12015/1 and MC_UU_00006/1) and Cancer Research UK (C864/A14136). The genetics work in the EPIC-Norfolk study was funded by the Medical Research Council (MC_PC_13048). We are grateful to all the participants who have been part of the project and to the many members of the study teams at the University of Cambridge who have enabled this research. Indiana Biobank was made possible, in part, with support from the Indiana Clinical and Translational Sciences Institute funded, in part by Award Number UL1TR002529 from the National Institutes of Health, National Center for Advancing Translational Sciences, Clinical and Translational Sciences Award, and the National Center for Research Resources, Construction grant number RR020128 and the Lilly Endowment. The content is solely the responsibility of the authors and does not necessarily represent the official views of the National Institutes of Health. The authors acknowledge the Indiana University Pervasive Technology Institute for providing [HPC (Big Red II, Karst, Carbonate), visualization, database, storage, or consulting] resources that have contributed to the research results reported within this paper. BWHHS is supported by funding from the British Heart Foundation and the Department of Health Policy Research Programme (England). PBM acknowledges the support of the National Institute for Health and Care Research Barts Biomedical Research Centre (NIHR203330). APK is supported by a UK Research and Innovation Future Leaders Fellowship, an Alcon Research Institute Young Investigator Award and a Lister Institute for Preventive Medicine Award. This research was supported by the NIHR Biomedical Research Centre at Moorfields Eye Hospital and the UCL Institute of Ophthalmology.

