## Supplementary Materials for "Utilizing multimodal AI to improve genetic analyses of cardiovascular traits"

immediate

### Supplementary Notes

#### Dataset acknowledgment

#### M-REGLE model architecture (ECG + PPG)

Encoder

| Layer (type) | Output Shape | Param # |
|---|---|---|
| vae_encoder_input (InputLayer) | [(None, 700, 1)] | 0 |
| vae_encoder_conv1 (Conv1D) | (None, 700, 16) | 128 |
| leaky_re_lu (LeakyReLU) | (None, 700, 16) | 0 |
| vae_encoder_pooling1 (MaxPooling1D) | (None, 350, 16) | 0 |
| vae_encoder_conv2 (Conv1D) | (None, 350, 16) | 1808 |
| leaky_re_lu_1 (LeakyReLU) | (None, 350, 16) | 0 |
| vae_encoder_pooling2 (MaxPooling1D) | (None, 175, 16) | 0 |
| vae_encoder_conv3 (Conv1D) | (None, 175, 16) | 1808 |
| leaky_re_lu_2 (LeakyReLU) | (None, 175, 16) | 0 |
| vae_encoder_pooling3 (MaxPooling1D) | (None, 88, 16) | 0 |
| vae_encoder_conv4 (Conv1D) | (None, 88, 16) | 1808 |
| leaky_re_lu_3 (LeakyReLU) | (None, 88, 16) | 0 |
| vae_encoder_pooling4 (MaxPooling1D) | (None, 44, 16) | 0 |
| vae_encoder_conv5 (Conv1D) | (None, 44, 16) | 1808 |
| leaky_re_lu_4 (LeakyReLU) | (None, 44, 16) | 0 |
| vae_encoder_pooling5 (MaxPooling1D) | (None, 22, 16) | 0 |
| vae_encoder_flatten (Flatten) | (None, 352) | 0 |
| vae_encoder_dense1 (Dense) | (None, 64) | 22592 |
| vae_encoder_dense2 (Dense) | (None, 32) | 2080 |
| z_mean (Dense) | (None, 12) | 396 |
| z_log_var (Dense) | (None, 12) | 396 |
| gaussian_sampling (GaussianSampling) | (None, 12) | 0 |

Total params: 32824

Trainable params: 32824

Decoder

| Layer (type) | Output Shape | Param # |
| --- | --- | --- |
| decoder_input (InputLayer) | [(None, 12)] | 0 |
| decoder_dense1 (Dense) | (None, 32) | 416 |
| decoder_dense2 (Dense) | (None, 64) | 2112 |
| decoder_dense3 (Dense) | (None, 352) | 22880 |
| reshape (Reshape) | (None, 22, 16) | 0 |
| decoder_upsample1 (UpSampling1D) | (None, 44, 16) | 0 |
| decoder_trans_conv1 (Conv1DTranspose) | (None, 44, 16) | 1808 |
| leaky_re_lu_5 (LeakyReLU) | (None, 44, 16) | 0 |
| decoder_upsample2 (UpSampling1D) | (None, 88, 16) | 0 |
| decoder_trans_conv2 (Conv1DTranspose) | (None, 88, 16) | 1808 |
| leaky_re_lu_6 (LeakyReLU) | (None, 88, 16) | 0 |
| decoder_upsample3 (UpSampling1D) | (None, 176, 16) | 0 |
| decoder_trans_conv3 (Conv1DTranspose) | (None, 176, 16) | 1808 |
| leaky_re_lu_7 (LeakyReLU) | (None, 176, 16) | 0 |
| decoder_upsample4 (UpSampling1D) | (None, 352, 16) | 0 |
| decoder_trans_conv4 (Conv1DTranspose) | (None, 352, 16) | 1808 |
| leaky_re_lu_8 (LeakyReLU) | (None, 352, 16) | 0 |
| decoder_upsample5 (UpSampling1D) | (None, 704, 16) | 0 |
| decoder_trans_conv5 (Conv1DTranspose) | (None, 704, 1) | 113 |
| leaky_re_lu_9 (LeakyReLU) | (None, 704, 1) | 0 |
| cropping1d (Cropping1D) | (None, 700, 1) | 0 |

Total params: 32753

Trainable params: 32753

#### M-REGLE model architecture (12 Lead ECG)

Encoder

| Layer (type) | Output Shape | Param # |
|---|---|---|
| vae_encoder_input (InputLayer) | [(None, 600, 12)] | 0 |
| vae_encoder_conv1 (Conv1D) | (None, 600, 32) | 1184 |
| leaky_re_lu (LeakyReLU) | (None, 600, 32) | 0 |
| vae_encoder_pooling1 (MaxPooling1D) | (None, 300, 32) | 0 |
| vae_encoder_conv2 (Conv1D) | (None, 300, 32) | 3104 |
| leaky_re_lu_1 (LeakyReLU) | (None, 300, 32) | 0 |
| vae_encoder_pooling2 (MaxPooling1D) | (None, 150, 32) | 0 |
| vae_encoder_conv3 (Conv1D) | (None, 150, 32) | 3104 |
| leaky_re_lu_2 (LeakyReLU) | (None, 150, 32) | 0 |
| vae_encoder_pooling3 (MaxPooling1D) | (None, 75, 32) | 0 |
| vae_encoder_flatten (Flatten) | (None, 2400) | 0 |
| vae_encoder_dense1 (Dense) | (None, 512) | 1229312 |
| z_mean (Dense) | (None, 96) | 49248 |
| z_log_var (Dense) | (None, 96) | 49248 |
| gaussian_sampling (GaussianSampling) | (None, 96) | 0 |

Total params: 1335200

Trainable params: 1335200

Decoder

| Layer (type) | Output Shape | Param # |
|---|---|---|
| decoder_input (InputLayer) | [(None, 96)] | 0 |
| decoder_dense1 (Dense) | (None, 512) | 49664 |
| decoder_dense2 (Dense) | (None, 2400) | 1231200 |
| reshape (Reshape) | (None, 75, 32) | 0 |
| decoder_upsample1 (UpSampling1D) | (None, 150, 32) | 0 |
| decoder_trans_conv1 (Conv1DTranspose) | (None, 150, 32) | 3104 |
| leaky_re_lu_3 (LeakyReLU) | (None, 150, 32) | 0 |
| decoder_upsample2 (UpSampling1D) | (None, 300, 32) | 0 |
| decoder_trans_conv2 (Conv1DTranspose) | (None, 300, 32) | 3104 |
| leaky_re_lu_4 (LeakyReLU) | (None, 300, 32) | 0 |
| decoder_upsample3 (UpSampling1D) | (None, 600, 32) | 0 |
| decoder_trans_conv3 (Conv1DTranspose) | (None, 600, 12) | 1164 |
| leaky_re_lu_5 (LeakyReLU) | (None, 600, 12) | 0 |
| cropping1d (Cropping1D) | (None, 600, 12) | 0 |

Total params: 1288236

Trainable params: 1288236

#### Supplementary Figures

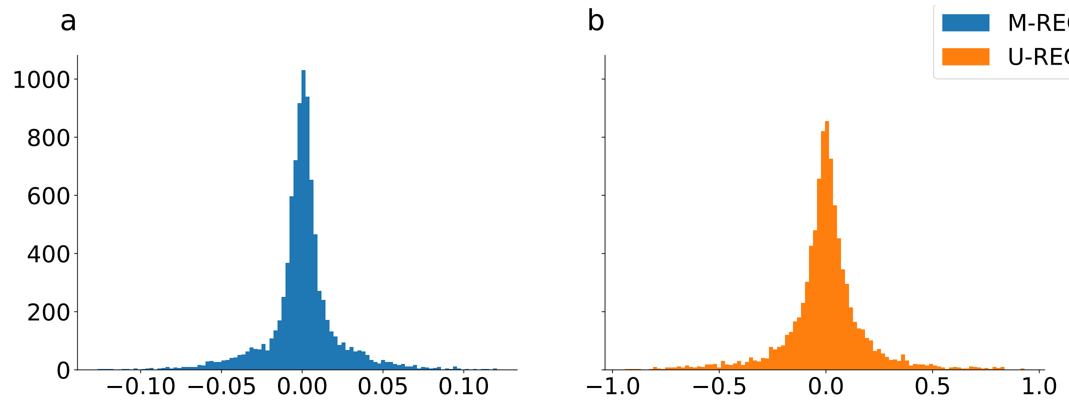

Supplementary Figure 1: **Correlation coefficients (non-diagonal) histogram of M-REGLE and U-REGLE embeddings of 12-lead ECG.** a) Histogram of non-diagonal correlation coefficient matrices of M-REGLE 12-lead ECG embeddings. Most of the non-diagonal coefficients fall in [-0.1,0.1], which shows that the coordinates of M-REGLE embeddings are mostly orthogonalized. b) Histogram of non-diagonal correlation coefficient matrices of U-REGLE 12-lead ECG embeddings.

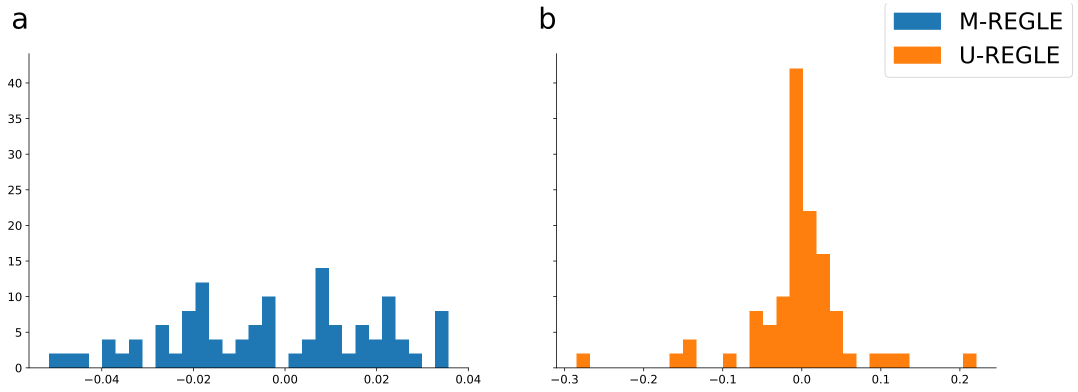

Supplementary Figure 2: **Correlation coefficients (non-diagonal) histogram of M-REGLE and U-REGLE embeddings of ECG lead I + PPG.** a) Histogram of non-diagonal correlation coefficient matrices of M-REGLE ECG lead I + PPG embeddings. Most of the non-diagonal coefficients fall in [-0.05,0.05], which shows that the coordinates of M-REGLE embeddings are mostly orthogonalized. b) Histogram of non-diagonal correlation coefficient matrices of U-REGLE ECG lead I + PPG embeddings.

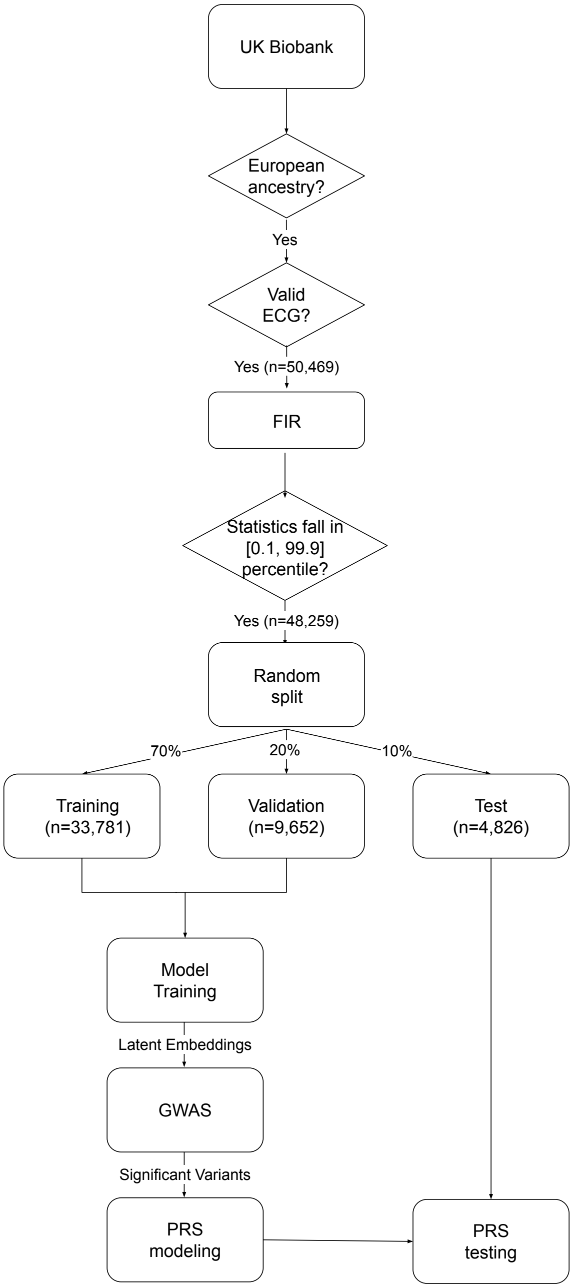

Supplementary Figure 3: **An overview of UK Biobank 12-lead ECG used in this study** Our initial dataset consists of all European-ancestry in UK Biobank (n=435,766). We considered all individuals with valid 12-lead ECG waveform at instance 2 (n=50,469). After the preprocessing and quality control steps, we split the dataset to training (70%) validation (20%) and test (10%) sets. We use all individuals in training and validation sets for GWAS analysis, and test set for reporting the PRS results.

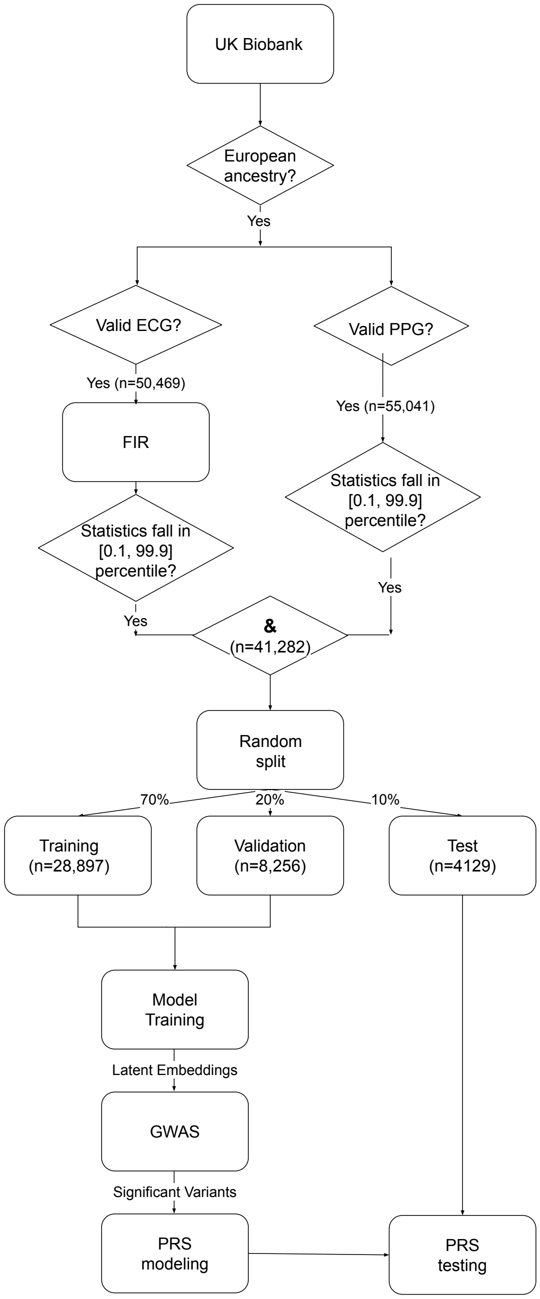

Supplementary Figure 4: **An overview of UK Biobank ECG lead I and PPG used in this study** Our initial dataset consists of all European-ancestry in UK Biobank (n=435,766). We considered all individuals with valid ECG lead I (n=50,469) and PPG (n=55,041) waveform at instance 2. After the preprocessing and quality control steps, we get all the individuals with both ECG and PPG qualified. We split the dataset to training (70%) validation (20%) and test (10%) sets. We use all individuals in training and validation sets for GWAS analysis, and test set for reporting the PRS results.

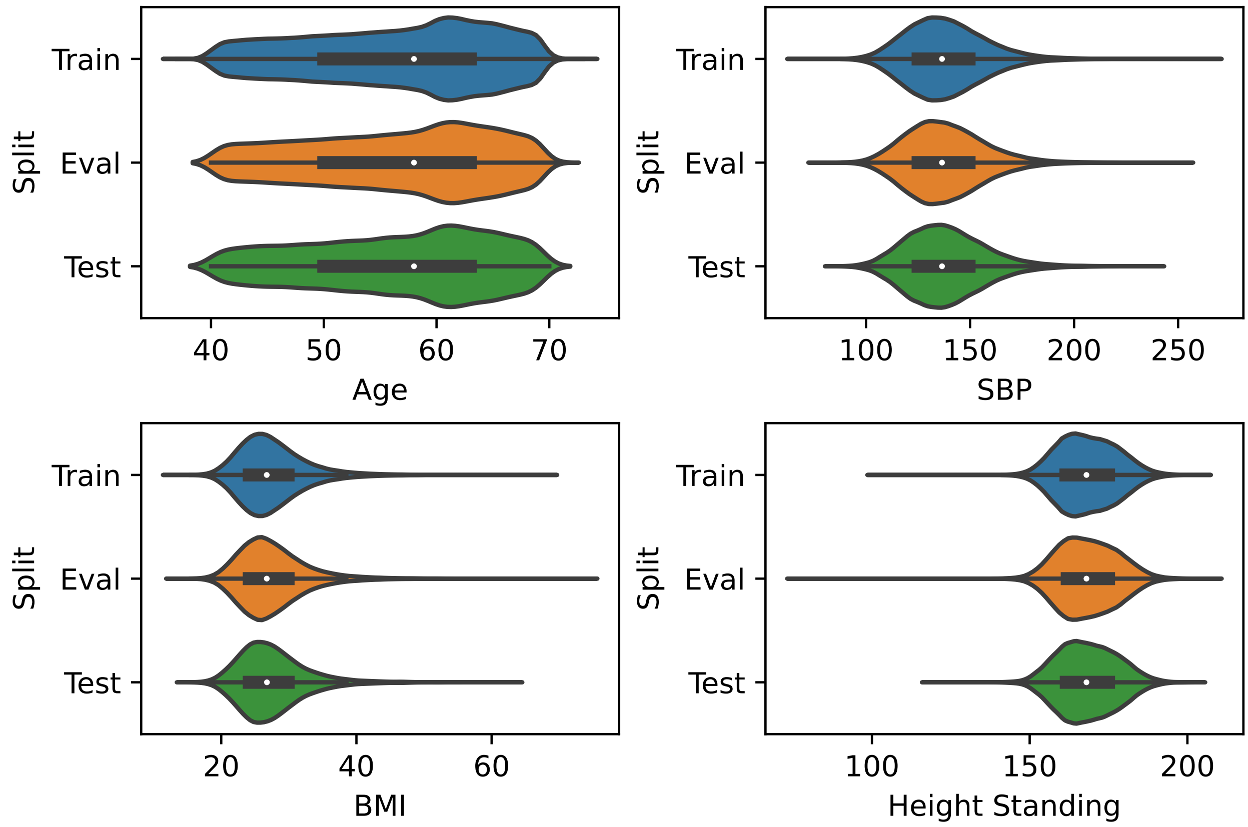

Supplementary Figure 5: **UK Biobank phenotype distribution for different data splits** We plot the age, SBP (systolic blood pressure), BMI (body mass index), and Height standing for train, eval, and test datasets in UKB. We observed that in most cases all datasets tend to have similar phenotypic distribution.

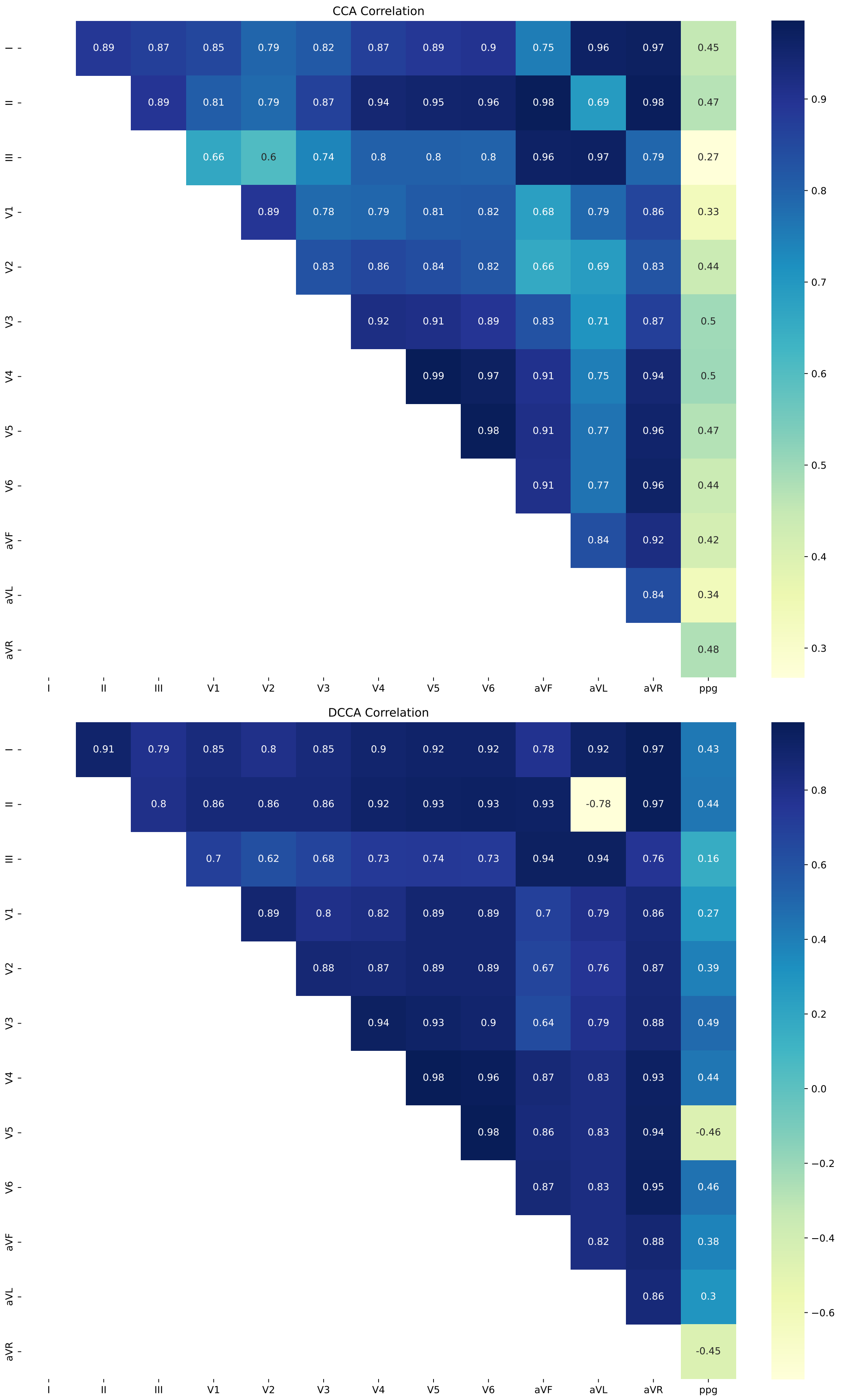

Supplementary Figure 6: **CCA and DCCA between 12 ECG leads and PPG in UK Biobank.** We plotted the mean correlation obtained from CCA and DCCA. The standard error of correlation and numerical values see Supplementary Table 2.

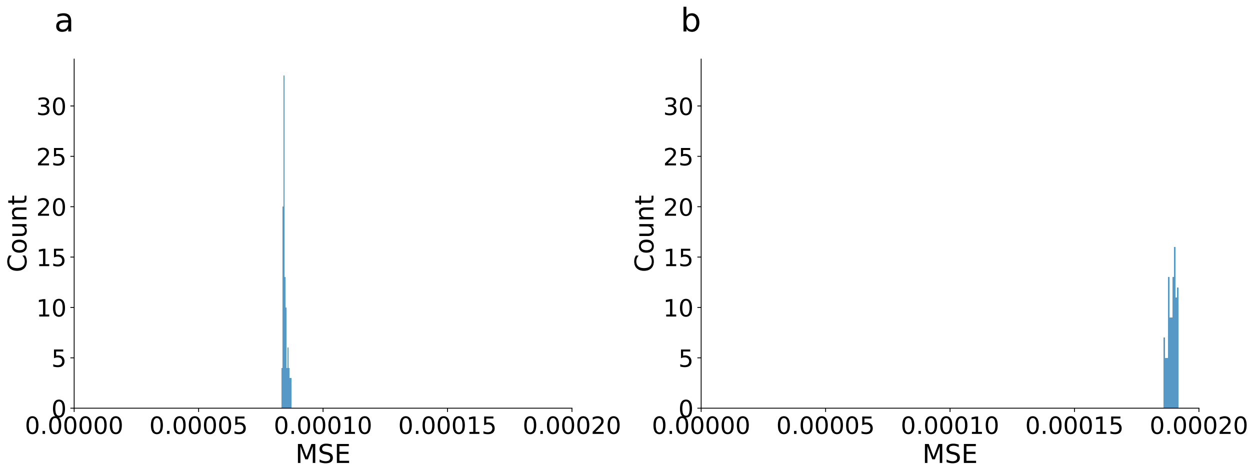

Supplementary Figure 7: **M-REGLE's performance is robust against random seeds.** a) Histogram of MSEs of 12-lead ECG models (96 latent dimensions) trained on 100 different random seeds. b) Histogram of MSEs of ECGPPG models (12 latent dimensions) trained on 100 different random seeds.

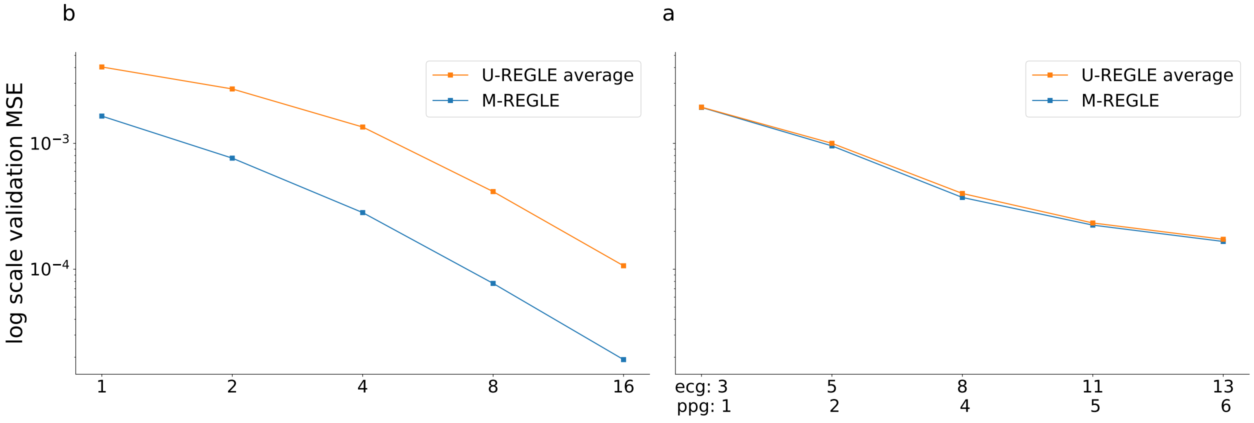

Supplementary Figure 8: **Reconstruction errors of unimodal modal PCA and multimodal PCA across different PC numbers.** a) Plot of log scale validation reconstruction loss of unimodal modal PCA and multimodal PCA across different PC numbers on Lead I ECG + PPG data setting.b) Plot of log scale validation reconstruction loss of unimodal modal PCA and multimodal PCA across different PC numbers on 12-lead ECG data setting

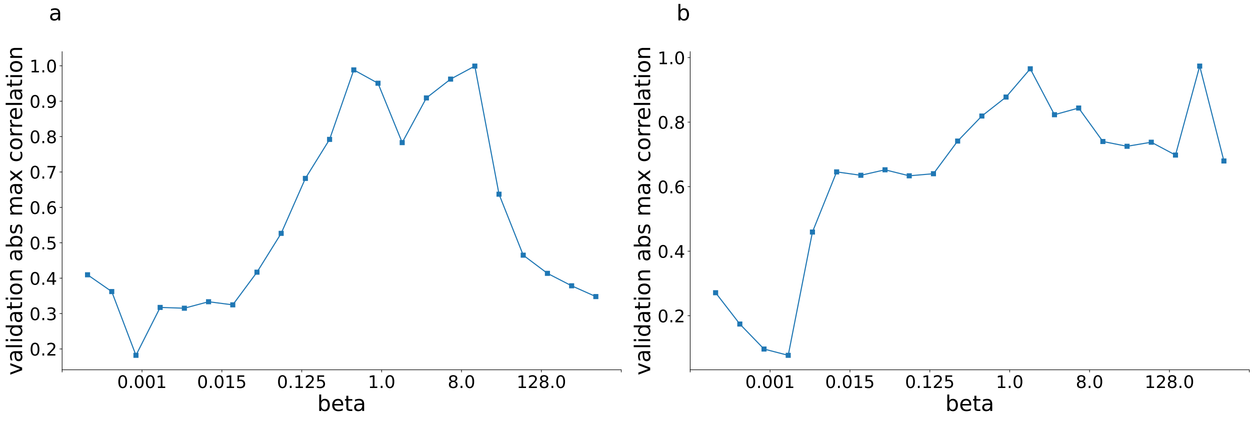

Supplementary Figure 9: **Plot of maximum absolute non-diagonal correlation coefficients of M-REGLE lower embeddings when training with different beta values.** a) Maximum absolute non-diagonal correlation coefficients VS beta plot of 12-lead ECG, b) Maximum absolute non-diagonal correlation coefficients VS beta plot of ECG+PPG.

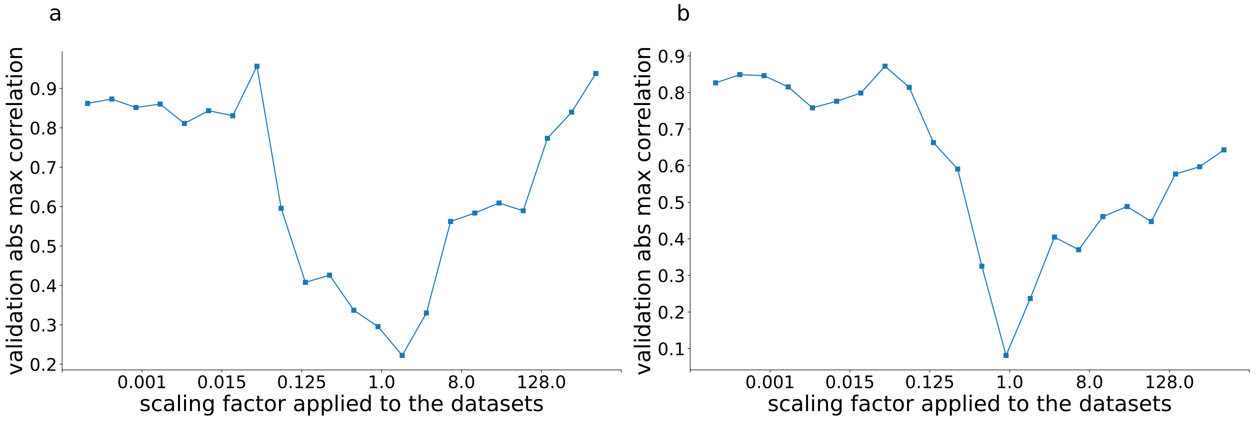

Supplementary Figure 10: **Plot of maximum absolute non-diagonal correlation coefficients of M-REGLE lower embeddings when different scaling factors applied to the datasets.** a) Maximum absolute non-diagonal correlation coefficients VS scaling factor plot of 12-lead ECG, b) Maximum absolute non-diagonal correlation coefficients VS scaling factor plot of ECG+PPG.

#### Supplementary Tables

| | Train | Eval | Test |
|---|---|---|---|
| Myocardial Infarction | 0.0593 | 0.0599 | 0.0585 |
| Afib | 0.0722 | 0.0729 | 0.0709 |
| T2D | 0.0836 | 0.0825 | 0.0828 |
| Cardiovascular Disease | 0.4149 | 0.4116 | 0.4150 |
| Hypertension | 0.3941 | 0.3909 | 0.3942 |
| Ever smoked | 0.6031 | 0.6013 | 0.6008 |

Supplementary Table 1: **UK Biobank binary phenotypes prevalence for different data splits.**

See the attached Excel table.

Supplementary Table 2: **CCA and DCCA between 12 ECG leads and PPG in UK Biobank.**

| # ECG Latent Dim. | ECG variance explained | # PPG Latent Dim. | PPG variance explained |
|---|---|---|---|
| 3 | 0.6807 | 1 | 0.6728 |
| 5 | 0.8353 | 2 | 0.8370 |
| 8 | 0.9311 | 4 | 0.9496 |
| 11 | 0.9616 | 5 | 0.9660 |
| 13 | 0.9716 | 6 | 0.9763 |
| 17 | 0.9834 | 7 | 0.9848 |
| 22 | 0.9902 | 8 | 0.9912 |

Supplementary Table 3: **Latent dimension number and variance expained through PCA for Lead I ECG and PPG.**

| Latent Dim. per channel | Multimodal | Unimodal |
|---|---|---|
| 1 | 1.00e-03 ± 6.77e-07 | 2.32e-03 ± 1.43e-06 |
| 2 | 4.13e-04 ± 2.55e-07 | 1.46e-03 ± 8.46e-07 |
| 4 | 1.58e-04 ± 1.68e-07 | 7.22e-04 ± 3.26e-07 |
| 8 | 7.32e-05 ± 8.40e-08 | 2.67e-04 ± 1.62e-07 |
| 16 | 6.41e-05 ± 7.18e-08 | 1.46e-04 ± 9.01e-08 |

Supplementary Table 4: **Comparison of multimodal VAE MSE and an average of all unimodal modal VAE MSEs across different latent dimension numbers on 12 lead ECG dataset.** Latent Dim stands for latent dimension.

| Latent Dim. per channel | Multimodal | Unimodal |
|---|---|---|
| 1 | 1.60e-03 ± 1.26e-06 | 3.88e-03 ± 2.60e-06 |
| 2 | 7.63e-04 ± 6.12e-07 | 2.62e-03 ± 1.80e-06 |
| 4 | 2.88e-04 ± 2.74e-07 | 1.33e-03 ± 8.02e-07 |
| 8 | 8.50e-05 ± 1.15e-07 | 4.38e-04 ± 3.44e-07 |
| 16 | 2.46e-05 ± 4.42e-08 | 1.17e-04 ± 1.49e-07 |

Supplementary Table 5: **Comparison of multimodal PCA MSE and an average of all unimodal modal PCA MSEs across different PC numbers on 12 lead ECG dataset.**

| Latent Dim. per channel | Multimodal | Unimodal |
|---|---|---|
| 1 | 1.03e-03 | 2.69e-03 |
| 2 | 4.32e-04 | 1.71e-03 |
| 4 | 1.64e-04 | 8.08e-04 |
| 8 | 7.71e-05 | 2.45e-04 |
| 16 | 4.70e-05 | 7.97e-05 |

Supplementary Table 6: **Comparison of multimodal CAE MSE and an average of all unimodal modal CAE MSEs across different latent dimension numbers on 12 lead ECG dataset.** Latent Dim stands for latent dimension.

| Variance explained | Multimodal | Unimodal |
|---|---|---|
| 70% | 9.49e-04 ± 1.98e-06 | 1.07e-03 ± 2.26e-06 |
| 83% | 5.09e-04 ± 8.86e-07 | 5.44e-04 ± 9.42e-07 |
| 93%-94% | 2.08e-04 ± 3.82e-07 | 2.60e-04 ± 4.46e-07 |
| 96% | 1.27e-04 ± 2.60e-07 | 1.95e-04 ± 3.36e-07 |
| 97% | 1.02e-04 ± 1.90e-07 | 1.86e-04 ± 3.01e-07 |

Supplementary Table 7: **Comparison of multimodal VAE MSE and an average of all unimodal modal VAE MSEs across different latent dimension numbers on lead I ECG + PPG dataset.**

| Variance explained | Multimodal | Unimodal |
|---|---|---|
| 70% | 1.93e-03 ± 4.64e-06 | 1.94e-03 ± 4.65e-06 |
| 83% | 9.54e-04 ± 2.78e-06 | 1.00e-03 ± 2.37e-06 |
| 93%-94% | 3.71e-04 ± 1.13e-06 | 3.99e-04 ± 1.29e-06 |
| 96% | 2.24e-04 ± 8.68e-07 | 2.33e-04 ± 8.66e-07 |
| 97% | 1.66e-04 ± 7.55e-07 | 1.73e-04 ± 7.84e-07 |

Supplementary Table 8: **Comparison of multimodal PCA MSE and an average of all unimodal modal PCA MSEs across different latent dimension numbers on lead I ECG + PPG dataset.**

| Variance explained | Multimodal | Unimodal |
|---|---|---|
| 70% | 9.72e-04 | 9.43e-04 |
| 83% | 5.48e-04 | 4.99e-04 |
| 93%-94% | 1.90e-04 | 1.93e-04 |
| 96% | 1.16e-04 | 1.14e-04 |
| 97% | 8.61e-05 | 8.47e-05 |

Supplementary Table 9: **Comparison of multimodal CAE MSE and an average of all unimodal modal CAE MSEs across different latent dimension numbers on lead I ECG + PPG dataset.**

See the attached Excel table.

Supplementary Table 10: **M-REGLE phenotypic correlation with UKB phenotypes.**

See the attached Excel table.

Supplementary Table 11: **Phenotype prediction metrics by 12-lead ECG M-REGLE & U-REGLE embeddings.**

See the attached Excel table.

Supplementary Table 12: **Paired test of phenotype prediction metrics by 12-lead ECG M-REGLE & U-REGLE embeddings.**

See the attached Excel table.

Supplementary Table 13: **Phenotype prediction metrics by ECG lead I + PPG M-REGLE & U-REGLE embeddings.**

See the attached Excel table.

Supplementary Table 14: **Paired test of phenotype prediction metrics by ECG lead I + PPG M-REGLE & U-REGLE embeddings.**

See the attached Excel table.

Supplementary Table 15: **LDSC heritability and intercept obtained for 96 embeddings obtained from 12-lead ECG.**

See the attached Excel table.

Supplementary Table 16: **M-REGLE 12 lead ECG GWS loci.** CHR, chromosome; POS, base-pair variant position; EA, effect allele; NEA, non-effect allele; SRC, imputed or genotyped variant; INFO, imputation INFO score (set to 1 for genotyped variants); GWAS p-value. GENE_CONTEXT, genomic context of the variant. Notation for gene context:

- Overlapping gene(s):
  - `[A]`: variant overlaps gene `A`
  - `[A,B]`: variant overlaps genes `A` and `B`
- Downstream genes:
  - `[]A`: variant position is $0 < p \leq 10^3$ bp upstream of closest downstream gene `A`
  - `[]-A`: variant position is $10^3 < p \leq 10^4$ bp upstream of closest downstream gene `A`
  - `[]-A`: variant position is $10^4 < p \leq 10^5$ bp upstream of closest downstream gene `A`
  - `[]--A`: variant position is $10^5 < p \leq 10^6$ bp upstream of closest downstream gene `A`
  - `[]`: closest downstream gene is further than $10^6$ bp
- Upstream genes: mirrors downstream gene notation, e.g., `B-[]` means variant position is $10^3 < p \leq 10^4$ bp downstream of closest gene `B`.

| Method | Num Discovered Hits | Num Discovered Loci |
|---|---|---|
| PCA-unimodal | 167 (148, 88.62%) | 104 (94, 90.38%) |
| PCA-multimodal | 215 (191, 88.84%) | 122 (108, 88.52%) |
| CAE-unimodal | 188 (166, 88.30%) | 114 (103, 90.35%) |
| CAE-multimodal | 227 (203, 89.43%) | 131 (116, 88.55%) |
| VAE-unimodal (U-REGLE) | 200 (182, 91.00%) | 119 (107, 89.92%) |
| VAE-multimodal (M-REGLE) | 262 (231, 88.17%) | 142 (122, 85.92%) |

Supplementary Table 17: **Comparison of rediscovered hits and loci by different models and learning methods on 12-lead ECG.** The numbers in each parenthesis are the number and percentage of rediscovered hits/loci in GWAS catalog.

| Method | Modality | $E[\chi^2]$ | se($E[\chi^2]$) | CI |
|---|---|---|---|---|
| PCA | unimodal | 71.54 | 1.38 | (68.85, 74.3) |
| PCA | multimodal | 90.98 | 1.68 | (87.74, 94.3) |
| CAE | unimodal | 88.22 | 1.77 | (84.84, 91.77) |
| CAE | multimodal | 99.24 | 1.93 | (95.53, 103.05) |
| VAE | unimodal | 92.22 | 1.82 | (88.75, 95.84) |
| VAE | multimodal | 112.48 | 2.06 | (108.67, 116.76) |

Supplementary Table 18: **Comparison of expected Chi-Square statistics of GWAS Catalog variants by different models and learning methods on Lead I ECG + PPG dataset.**

| Modality | Merge | Model | Num loci | Num cardio enrichment |
|---|---|---|---|---|
| ECG | Single | PCA | 104 | 47 |
| ECG | Single | CAE | 114 | 48 |
| ECG | Single | VAE | 119 | 68 |
| ECG | Multi | PCA | 122 | 48 |
| ECG | Multi | CAE | 131 | 63 |
| ECG | Multi | VAE | 142 | 61 |
| ECG+PPG | Single | PCA | 44 | 35 |
| ECG+PPG | Single | CAE | 54 | 57 |
| ECG+PPG | Single | VAE | 54 | 61 |
| ECG+PPG | Multi | PCA | 45 | 32 |
| ECG+PPG | Multi | CAE | 59 | 50 |
| ECG+PPG | Multi | VAE | 61 | 69 |

Supplementary Table 19: **Comparison of GREAT enriched cardiovascular term counts by different models and learning methods.**

| $Model_1$ | $Model_2$ | Stronger model | Nominal P-value |
|---|---|---|---|
| ECG Single PCA | ECG Single CAE | - | 0.062 |
| ECG Single PCA | ECG Single VAE | Single VAE | 3.0e-41 |
| ECG Single CAE | ECG Single VAE | Single VAE | 2.3e-43 |
| ECG Single PCA | ECG Multi PCA | Multi PCA | 3.9e-13 |
| ECG Single CAE | ECG Multi CAE | Multi CAE | 1.1e-29 |
| ECG Single VAE | ECG Multi VAE | Multi VAE | 0.022 |
| ECG Multi PCA | ECG Multi CAE | Multi CAE | 1.5e-11 |
| ECG Multi PCA | ECG Multi VAE | Multi VAE | 2.0e-19 |
| ECG Multi CAE | ECG Multi VAE | Multi VAE | 8.2e-6 |
| ECG+PPG Single PCA | ECG+PPG Single CAE | Single CAE | 1.3e-13 |
| ECG+PPG Single PCA | ECG+PPG Single VAE | Single VAE | 1.4e-5 |
| ECG+PPG Single CAE | ECG+PPG Single VAE | Single CAE | 8.9e-5 |
| ECG+PPG Single PCA | ECG+PPG Multi PCA | Single PCA | 1.1e-11 |
| ECG+PPG Single CAE | ECG+PPG Multi CAE | Multi CAE | 1.4e-5 |
| ECG+PPG Single VAE | ECG+PPG Multi VAE | Multi VAE | 1.9e-14 |
| ECG+PPG Multi PCA | ECG+PPG Multi CAE | Multi CAE | 4.2e-20 |
| ECG+PPG Multi PCA | ECG+PPG Multi VAE | Multi VAE | 3.7e-32 |
| ECG+PPG Multi CAE | ECG+PPG Multi VAE | Multi VAE | 5.0e-14 |

Supplementary Table 20: **Comparison of significance of significantly enriched cardiovascular term p-values by different models and learning methods.** P-values were computed by two-sided paired T-test on -$\log_{10}$-transformed ontology term nominal p-values.

See the attached Excel table.

Supplementary Table 21: **LDSC heritability and intercept obtained for 12 embeddings obtained from ECG lead I and PPG.**

See the attached Excel table.

Supplementary Table 22: **M-REGLE lead I ECG and PPG loci.** CHR, chromosome; POS, base-pair variant position; EA, effect allele; NEA, non-effect allele; SRC, imputed or genotyped variant; INFO, imputation INFO score (set to 1 for genotyped variants); GWAS p-value. GENE_CONTEXT, genomic context of the variant. Notation for gene context:

- Overlapping gene(s):
  - `[A]`: variant overlaps gene `A`
  - `[A,B]`: variant overlaps genes `A` and `B`
- Downstream genes:
  - `[]A`: variant position is $0 < p \leq 10^3$ bp upstream of closest downstream gene `A`
  - `[]-A`: variant position is $10^3 < p \leq 10^4$ bp upstream of closest downstream gene `A`
  - `[]-A`: variant position is $10^4 < p \leq 10^5$ bp upstream of closest downstream gene `A`
  - `[]--A`: variant position is $10^5 < p \leq 10^6$ bp upstream of closest downstream gene `A`
  - `[]`: closest downstream gene is further than $10^6$ bp
- Upstream genes: mirrors downstream gene notation, e.g., `B-[]` means variant position is $10^3 < p \leq 10^4$ bp downstream of closest gene `B`.

| Method | Num Discovered Hits | Num Discovered Loci |
|---|---|---|
| PCA-unimodal | 69 (67, 97.10%) | 44 (43, 97.73%) |
| PCA-multimodal | 69 (68, 98.55%) | 45 (45, 100.00%) |
| CAE-unimodal | 77 (74, 96.10%) | 54 (52, 96.30%) |
| CAE-multimodal | 93 (89, 95.70%) | 59 (57, 96.61%) |
| VAE-unimodal (U-REGLE) | 89 (85, 95.51%) | 54 (52, 96.30%) |
| VAE-multimodal (M-REGLE) | 103 (99, 96.12%) | 61 (59, 96.72%) |

Supplementary Table 23: **Comparison of rediscovered hits and loci by different models and learning methods on Lead I ECG + PPG dataset.** The numbers in each parenthesis are the number and percentage of rediscovered hits/loci in GWAS catalog.

| Method | Modality | $E[\chi^2]$ | se($E[\chi^2]$) | CI |
|---|---|---|---|---|
| pca | unimodal | 33.12 | 0.68 | (31.82, 34.48) |
| pca | multimodal | 33.23 | 0.69 | (31.95, 34.66) |
| cae | unimodal | 37.93 | 0.73 | (36.51, 39.41) |
| cae | multimodal | 46.05 | 0.89 | (44.34, 47.85) |
| vae | unimodal | 44.37 | 0.88 | (42.71, 46.16) |
| vae | multimodal | 51.66 | 0.97 | (49.81, 53.62) |

Supplementary Table 24: **Comparison of expected Chi-Square statistics of GWAS Catalog variants by different models and learning methods on 12 lead ECG dataset.** CI: confidence interval.

See the attached Excel table.

Supplementary Table 25: **M-REGLE loci improves phenotypic prediction compared to unimodal loci.**

| Method | AUROC | AUPRC | Top 1% prevalence | Top 5% prevalence |
|---|---|---|---|---|
| M-REGLE | 0.55 (0.53-0.57) * | 0.22 (0.21-0.24) | 0.21 (0.10-0.34) | 0.24 (0.18-0.30) |
| U-REGLE | 0.53 (0.51-0.55) | 0.21 (0.20-0.23) | 0.27 (0.17-0.40) | 0.25 (0.19-0.31) |

Supplementary Table 26: **Comparison of M-REGLE vs U-REGLE Afib prediction in Indiana Biobank dataset.** Star (*) indicates M-REGLE is significantly better than U-REGLE based on paired bootstrap test.

| Method | AUROC | AUPRC | Top 1% prevalence | Top 5% prevalence |
|---|---|---|---|---|
| M-REGLE | 0.58 (0.56-0.59)* | 0.21 (0.20-0.22)* | 0.25 (0.19-0.32) | 0.24 (0.22-0.27)* |
| U-REGLE | 0.55 (0.54-0.56) | 0.19 (0.18-0.20) | 0.22 (0.16-0.29) | 0.20 (0.18-0.23) |

Supplementary Table 27: **Comparison of M-REGLE vs U-REGLE Afib prediction in EPIC-Norfolk dataset.** Star (*) indicates M-REGLE is significantly better than U-REGLE based on paired bootstrap test.

| Phenotype | Method | Pearson R |
|---|---|---|
| PPG pulse rate | M-REGLE | 0.10 (0.08-0.11) |
| PPG pulse rate | U-REGLE | 0.09 (0.08-0.10) |
| SBP | M-REGLE | 0.03 (0.01-0.04) |
| SBP | U-REGLE | 0.02 (0.00-0.03) |

Supplementary Table 28: **Comparison of M-REGLE vs U-REGLE PPG pulse rate, ECG QT interval, and SBP prediction in EPIC-Norfolk dataset.**

| Method | AUROC | AUPRC | Top 1% prevalence | Top 5% prevalence |
|---|---|---|---|---|
| M-REGLE | 0.55 (0.52-0.58) | 0.11 (0.09-0.13)* | 0.16 (0.08-0.28) | 0.16 (0.12-0.21)* |
| U-REGLE | 0.53 (0.50-0.56) | 0.10 (0.08-0.11) | 0.09 (0.03-0.22) | 0.09 (0.05-0.13) |

Supplementary Table 29: **Comparison of M-REGLE vs U-REGLE Afib prediction in BWHHS dataset.** Star (*) indicates M-REGLE is significantly better than U-REGLE based on paired bootstrap test.

| Phenotype | Method | Pearson R |
|---|---|---|
| QT interval | M-REGLE | 0.003 (-0.030-0.036) |
| QT interval | U-REGLE | -0.002 (-0.036-0.036) |
| PPG pulse rate | M-REGLE | 0.08 (0.046-0.115) |
| PPG pulse rate | U-REGLE | -0.020 (-0.054-0.014) |
| SBP | M-REGLE | 0.028 (-0.006-0.061) |
| SBP | U-REGLE | 0.034 (0.0003-0.066) |

Supplementary Table 30: **Comparison of M-REGLE vs U-REGLE PPG pulse rate, ECG QT interval, and SBP prediction in BWHHS dataset.**

See the attached Excel table.

Supplementary Table 31: **Overview of hyperparameter sweep ranges for M-REGLE and U-REGLE VAE models.**

See the attached Excel table.

Supplementary Table 32: **Overview of final hyperparameters used in M-REGLE and U-REGLE VAE models.**

| 12 lead ECG key words | lead I ECG + PPG key words |
|---|---|
| Electrocardio | Electrocardio |
| ECG | ECG |
| EKG | ECK |
| PR interval | PR interval |
| Ventricular rate | Ventricular rate |
| PP interval | PP interval |
| PQ interval | PQ interval |
| QRS duration | QRS duration |
| QT interval | QT interval |
| RR interval | RR interval |
| P axis | P axis |
| R axis | R axis |
| T axis | T axis |
| | Arterial stiffness |
| | Pulse wave |
| | Pulse waveform |
| | Pulse-wave |
| | Notch position |

Supplementary Table 33: **Regular expression key words used for search in GWAS catalog.**
